# Single-cell RNA sequencing reveals immunosuppressive pathways associated with metastatic breast cancer

**DOI:** 10.1101/2024.09.25.24314388

**Authors:** Furkan Ozmen, Tugba Y. Ozmen, Aysegul Ors, Mahnaz Janghorban, Matthew J. Rames, Xi Li, Fariba Behbod, Gordon B. Mills, Hisham Mohammed

## Abstract

Metastatic breast cancer remains largely incurable, and the mechanisms underlying the transition from primary to metastatic breast cancer remain elusive. We analyzed the complex landscape of primary and metastatic breast cancer using scRNA-seq data from twenty-three female patients with either primary or metastatic disease to elucidate the genetic and molecular mechanisms underlying changes in the metastatic tumor ecosystem. We identify specific subtypes of stromal and immune cells critical to forming a pro-tumor microenvironment in metastatic lesions, including CCL2+ macrophages, cytotoxic T cells with an exhausted gene signature, and FOXP3+ regulatory T cells. Analysis of cell-cell communication highlights a marked decrease in tumor-immune cell interactions in metastatic tissues, likely strengthening the immunosuppressive microenvironment. In contrast, primary breast cancer samples displayed increased activation of the TNF-α signaling pathway via NF-kB, indicating a potential therapeutic target. Our study comprehensively characterizes the transcriptional landscape encompassing primary and metastatic breast cancer.

## Introduction

Breast cancer, which remains the most prevalent cancer in women, is a diverse and complex disease with a wide range of clinical manifestations and outcomes. The shift from an early localized primary tumor to metastatic lesions in distant organs represents a pivotal moment in the clinical course and prognosis of the disease. Despite advances in early detection and treatment, progression to metastatic disease continues to pose a significant clinical challenge with an unfavorable prognosis^1^. Individuals diagnosed with localized breast cancer typically exhibit an overall survival rate exceeding 90%^2^. Conversely, the prognosis drastically declines when cancer progresses to distant metastasis, with survival rates plummeting to around 25%^3^. Therefore, understanding the complex mechanisms and differences between primary and metastatic breast cancer is essential for informing treatment approaches and improving patient outcomes.

Tumor metastasis requires a complex, orchestrated cascade involving the inherent characteristics of tumor cells, such as genetic mutations, and the intricate interplay between cancer cells and different cellular elements within the tumor microenvironment (TME). This dynamic interaction encompasses a range of participants, including immune cells, tumor-associated macrophages (TAMs) and lymphoid cells, cancer-associated fibroblasts (CAFs), and components of the extracellular matrix (ECM)^4–6^.

Understanding the complex interactions between different cellular components in the TME is crucial for comprehending the mechanisms of tumor initiation, progression to metastasis, and prognosis. Studies have uncovered mutational and transcriptional signatures that are more frequent in breast cancer metastases using bulk genomic sequencing methods^7,8^. However, genetic signatures arising from bulk sequencing cannot decipher the sources of observed differences or the dynamic interplay between the cell types shaping the metastatic microenvironment in advanced breast cancer.

Single-cell RNA sequencing (scRNA-seq) can reveal the distinct transcriptional profiles of individual malignant and non-malignant cells in the tumor ecosystem. This has enabled analysis of complex intra-tumoral heterogeneity among TME interactions in BCs, such as triple-negative breast cancer^9^. In particular, previous studies have suggested that FOXP3+ regulatory T cells (Tregs) in breast cancer may lead to immune tolerance and poorer overall survival^10^, while cytotoxic T cells with exhausted gene expression patterns might characterize an immunosuppressive TME^11^.While immune deregulations are undeniably a core component of the transition to metastatic disease, comprehensive transcriptomic profiling comparing the primary and metastatic breast cancer TME at single-cell resolution has only been applied to a limited number of cases and metastatic sites^12^.

Here, we conduct scRNA-seq analysis to deconvolve the TME landscape in primary and metastatic ER+ breast cancer. We elucidate distinct gene expression profiles of tumor cells in primary and metastatic breast cancer while identifying specific subtypes of stromal and immune cells that may collectively contribute to developing an immunosuppressive microenvironment within metastatic tumors. Our study provides an overview of the underlying functional landscape of primary and metastatic breast cancer cells while shedding light on the heterogeneity and transcriptomic TME patterns underpinning disease progression.

## Results

### Landscape of primary and metastatic breast cancer via scRNA-seq

Despite the limited survival of patients with metastatic breast cancer, little is known about the evolution that occurs between normal and malignant cells in the tumor ecosystem of primary and metastatic breast cancer. To investigate this issue, scRNA-seq was performed on an all-female patient cohort comprising individuals diagnosed with either primary (n=12) or metastatic ER+ breast cancer (n=11). Multiple metastatic sites, including the liver, bone, lymph nodes, mediastinum, adrenal gland, and skin, were sampled (Fig.1a). All patients were classified as estrogen receptor-positive (ER+) based on IHC analysis (Supp. Table 1).

After quality control, removal of batch effects, and principal component analysis, 56,384 single cells from primary breast cancer tissues and 42,813 single cells from metastatic breast cancer tissues, a total of 99,197 cells, were visualized using UMAP for downstream analysis. The cells were partitioned into fifty-four clusters, consisting of seven main cell types: malignant cells, myeloid cells, T cells, natural killer (NK) cells, B cells, endothelial cells, and fibroblasts (Fig. 1b, 1c). Each cell type was characterized using established gene expression markers^13–15^ (Fig. 1d). Additional details and corresponding references for the methodologies employed can be found in ’Methods’.

**Fig. 1.**
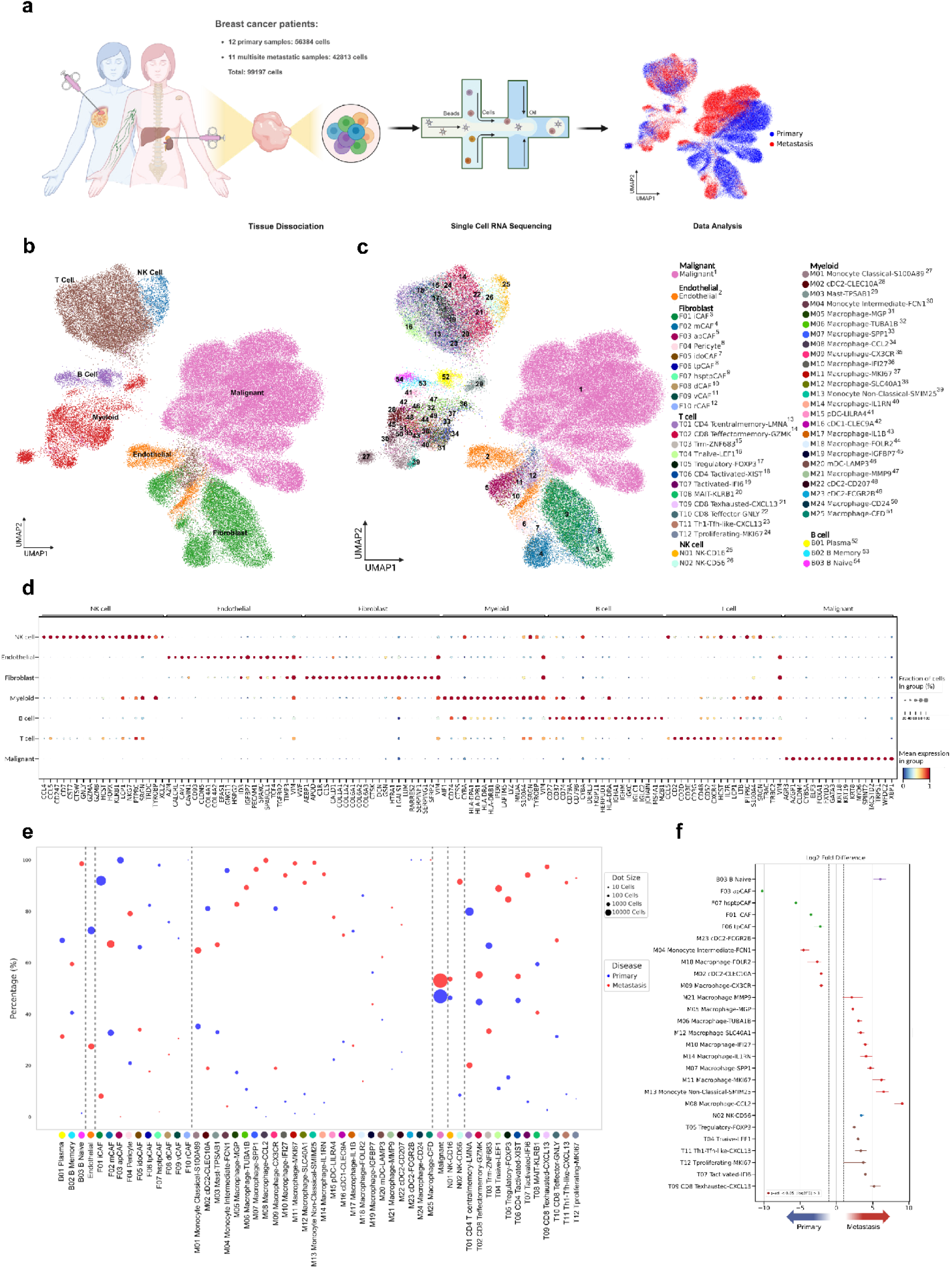
Single-Cell Landscape of Primary and Metastatic ER+ Breast Cancer: Characterization and Cellular Subtype Differences. **a.** Sample collection workflow and data analysis overview. (Created in BioRender. BioRender.com/y20n356) **b.** UMAP visualization of the unified cell map, consisting of 7 major cell types colored by clusters. **c.** UMAP visualization of the unified cell map, sub-segmenting by 54 minor cell types uniquely colored within the 7 major cell types. **d.** Dot plot representation of differentially expressed genes across the 7 major cell type clusters. **e.** Relative percentage of 54 minor cell types by metastatic status and corresponding cell number indicated by dot size. **f.** Significant changes in minor cell type proportions between primary and metastatic breast cancer

Copy number variation (CNV) profiles were determined using gene expression data and used to identify normal and malignant cells (see Methods). While both primary and metastatic samples exhibited the same main cell types, the proportions of each cell type varied widely between patients (Supp. Fig.1a, 1b). However, when we explored the differences in minor cell types (subtypes) within each group, we found a clear distinction in the proportion of cellular subtypes associated with primary and metastatic disease (Fig. 1e, 1f, Supp. Fig. 1c). Malignant epithelial cells were present in similar proportions in both primary and metastatic samples. In primary samples, FOLR2 and CXCR3 positive macrophages, which have been associated with a pro-inflammatory phenotype^16–19^, were predominantly observed. In contrast, macrophages positive for CCL2 and SPP1, which have been associated with a pro-tumorigenic subtype^20,21^, were more abundant in metastatic samples. Our observations highlight changes in the TME that are associated with primary and metastatic tumors, potentially underlying the transition toward the metastatic state.

### Genomic and phenotypic alterations within malignant cells

To identify cell types that exhibit high levels of gene expression heterogeneity, we performed differential gene expression analysis between patients for each major lineage. We found that malignant cells exhibited the most remarkable diversity of differentially expressed genes (DEGs), indicating pronounced transcriptional dynamics within these cellular populations across varying environmental conditions (Fig. 2a). To validate our malignant cell assignments and further define the tumor phenotype and clonal substructure, we estimated CNV using InferCNV^22^ and CaSpER^23^. T cells were used as a reference for each condition (primary/metastasis) (Fig. 2b, Supp. Fig. 2a, Supp. Document 1).

**Fig. 2.**
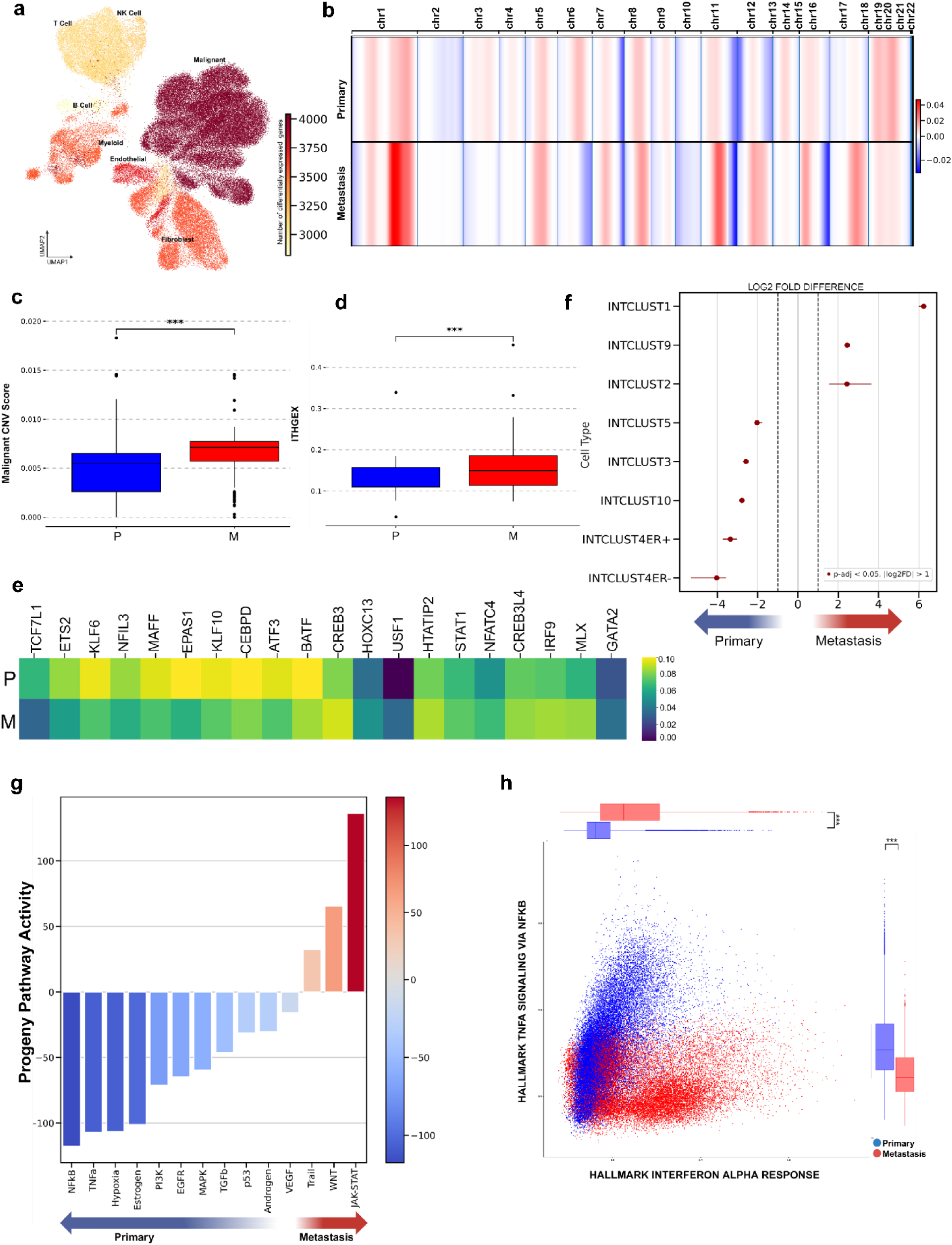
Genomic and Phenotypic Alterations in Malignant Cells of Primary vs. Metastatic Breast Cancer: Insights from CNV Analysis, Gene Regulatory Networks, and Integrative Clusters. **a.** UMAP visualization of the number of differentially expressed genes within all major cell type clusters. **b.** Chromosomal CNV alternation score by metastatic status, with copy number gain and copy number loss indicated by red and blue, respectively. **c.** CNV score comparison between primary and metastatic breast cancer malignant cells. Significance indicated as (***, p < 0.001). **d.** ITGEX score comparison between primary and metastatic breast cancer malignant cells. Significance indicated as (***, p < 0.001). **e.** Regulon activity scores by metastatic status within malignant cells. **f.** Significant (Log2 FC) integrative cluster differences between primary and metastatic malignant cells (p-adj < 0.05). **g.** Differential progeny pathway activity scores within malignant cells across metastatic status. **h.** Cellular states plot showing hallmark gene signature activity for Interferon Alpha response versus TNF-α signaling via NFkB in malignant cell populations.

CNV analysis revealed substantial copy number alterations in both primary and metastatic disease. Our analysis identified substantial inter-patient differences within the primary or metastatic group. We then compared the overall CNV structure in both groups and identified substantial inter-site differences. Notably, we observed significant variations in CNVs on chromosomes 1, 6, 11, 12, 16, and 17 when comparing primary and metastatic samples. These unique genomic alterations could potentially play a role in facilitating the transition to metastasis^24^.

To comprehend the distinct clonal structures between primary and metastatic breast cancer patients, we integrated CNVs from all tumor subpopulations per patient. We compared the overall pattern of copy number alterations across chromosomal arms. Our analysis led us to identify the top 25 CNVs within chromosomal arms that were specific to metastatic or primary subclones (Supp. Fig. 2b). In particular, CNVs in specific chromosomal regions, namely, chr7q34-q36, chr2p11-q11, chr16q13-q24, chr11q21-q25, chr12q13, chr7p22, and chr1q21-q44, were more frequent in the metastatic samples. Intriguingly, these regions encompass genes that have previously been associated with progression and aggressiveness of different cancer types, including ARNT, BIRC3, EIF2AK1, EIF2AK2, FANCA, HOXC11, KIAA1549, MSH2, MSH6, and MYCN^25–32^. These genes are associated with various aspects of cancer development and progression, including cell growth, proliferation, metabolism, and survival^25–32^. Elevated expression of several of the genes affected by these chromosomal alterations has also been reported to be associated with poorer survival rates in ER+ breast tumors^33^ (Supp. Fig. 3a).

We then calculated CNV scores for each cell (see Methods) using InferCNV, which represents the extent of copy number variations within a cell and reflects genomic instability. We found higher CNV scores in tumor cells from metastatic patients compared to primary breast samples (Fig. 2c). This finding is consistent with previous studies that have linked high CNV scores to poor prognosis in various types of cancer^34,35^. Besides intertumoral heterogeneity, intratumoral heterogeneity presents another significant challenge in accurately depicting the genomic landscape^36^. To further investigate intratumoral heterogeneity of copy number alterations, we used the SCEVAN^37^ algorithm to identify tumor sub-populations with different copy number alterations for each sample. Our observations indicate that metastatic tumors have a higher intratumoral heterogeneity gene expression score (ITHGEX)^38^ compared to primary tumors, consistent with metastatic breast cancer samples exhibiting higher levels of intratumoral heterogeneity (Fig. 2d). This is further evidenced by the existence of more tumor subclones within metastatic samples (Supp. Fig. 2b, Supp. Table 2).

### Gene regulatory signatures across primary and metastatic breast cancer

Next, we aimed to uncover the cellular processes potentially involved in primary and metastatic breast cancer by constructing lineage-specific gene regulatory networks (GRNs) based on transcription factor (TF) activity and their associated targets using SCENIC^39^. Through GRN analysis, we identified specific regulatory mechanisms in each significant cell type for primary and metastatic breast cancer. Our study revealed distinct expression patterns of the transcription factors associated with breast cancer pathophysiology (Fig. 2e).

Primary breast cancer malignant cells demonstrated higher regulon activities of transcription factors such as ETS2, EPAS1, BATF, NFIL3, TCF7L1, KLF6, MAFF, KLF10, CEBPD, and ATF3. These transcription factors are involved in various cellular functions implicated in tumor development, apoptosis, immune cell differentiation, energy metabolism, and regulation of essential signaling pathways^40–46^. For instance, ETS2 is known to regulate both tumor initiation and apoptosis^40^, while EPAS1 plays a crucial role in cellular responses to hypoxia^41^. BATF controls the differentiation of immune cells^42^, while NFIL3 is involved in energy metabolism and immune cell differentiation^43,44^. TCF7L1 mediates the Wnt signaling pathway^45^, and KLF6 modulates metabolism, immunity, and oncogenesis^46^.

In metastatic breast cancer, higher regulon activities of transcription factors, including HOXC13, GATA2, IRF9, MLX, CREB3L4, NFATC4, STAT1, HTATIP2, USF1, and CREB3, indicate their potential roles in facilitating cancer metastasis by influencing various biological processes. For instance, HOXC13 and GATA2 have been linked to a poor prognosis and aggressive phenotypes in a number of cancer lineages^47,48^. IRF9, an interferon response marker, is primarily known for its role in anti-viral immunity and has been linked to tumor growth and metastasis^49^. MLX coordinates lipid storage with metabolic gene expression regulation and is linked to poor prognosis^50^, whereas CREB3L4 is involved in unfolded protein response and has been associated with breast carcinoma progression^51^.

The differential activities of specific regulons in malignant cells reveal distinct mechanisms that could potentially be targeted in primary and metastatic tumor states.

### Metastatic breast cancer displays an enrichment for more aggressive Integrative Clusters

We scrutinized gene expression data and classified individual cells using Integrative Clusters (IntClust) derived from the METABRIC study^52^. The classification was based on raw expression data derived from the METABRIC study, using the top two hundred DEGs for each IntClust. To compute scores for each IntClust and classify each malignant cell, we employed the Cluster Independent Annotation (CIA) tool^53^. We performed a statistical proportion analysis for each cluster to determine whether any cell type was preferentially enriched or depleted for each method.

Malignant cells in primary tumors showed an increased presence of signatures associated with IntClust3, 4ER+, 4ER−, 5, and 10. IntClust3 is known for distinct patterns of chemosensitivity, and IntClust10 is associated with a stable probability of relapse-free cases among ER patients after five years^54^. In contrast, in metastatic malignant cells, we observed significant enrichment of signatures associated with IntClust1, 2, and 9. These three IntClust types are associated with more aggressive tumor behavior^52–54^. IntClust1 is characterized by late-recurring ER-positive genomic subgroups^54^, IntClust2 is associated with genomic alterations that contribute to its aggressive behavior^54^, and IntClust9 is distinguished by amplification of the MYC oncogene at 8q24 in 89% of tumors^54^ (Fig. 2f). Furthermore, we observed significant intratumor heterogeneity in the expression of IntClust types suggesting that tumors may contain cells with differing IntClust subtypes. The dominant IntClust types that represent the most frequent cellular subtypes varied for each sample, highlighting the complexity of the underlying cellular landscape (Supp. Fig. 3b).

### Primary breast cancer samples displayed increased activation of the TNF-α signaling pathway via NFkB

TNF-α and NFkB signaling pathway activities were elevated in malignant cells of primary breast cancer samples compared to those of metastatic samples. In contrast, we observed higher JAK-STAT pathway activity in metastatic samples compared to primary samples using Progeny pathway activity^55^ (Fig. 2g, 2h). We further investigated highly expressed genes from the NF-kB and JAK-STAT pathways in malignant cells from primary and metastatic samples. We found that many of the JAK-STAT-related genes enriched in metastatic malignant cells were also associated with interferon activity, while malignant cells from primary breast samples were enriched for genes associated positively with chemotaxis or immune regulation such as CCL20 and CXCL2^56,57^ (Supp. Fig. 3c, 3d, 3e). A similar trend was observed at the single-cell level, where malignant cells from primary samples were mostly in a cell state defined by higher TNF-α signaling via NF-KB, whereas malignant cells from metastatic samples were enriched in either hypoxia or interferon-alpha response cell states.

Given the importance of these pathways in regulating the immune response and altering the tumor microenvironment, we further investigated changes within non-malignant cells to assess potential deregulations in the immune microenvironment.

### Macrophage subtypes are polarized toward immunosuppression across metastatic breast cancer

Myeloid cells play pivotal roles in the TME. We identified twenty-five distinct myeloid subclusters in both primary and metastatic cancer types. These include fifteen subtypes of macrophages, six subtypes of dendritic cells (DCs), three subtypes of monocytes, and a single subtype of mast cells (Fig. 3a). Specific macrophage subtypes, including FOLR2-expressing macrophages, CX3CR-expressing macrophages, and FCN1- expressing monocytes, were notably enriched in primary tumors (Fig. 3b, 3c). FOLR2- positive macrophages in the tumor microenvironment have been observed to have increased communication with CD8+ T cells. This interaction is facilitated by the high expression of CXCL9 and CXCR3, which are closely associated with FOLR2^18,58^.

**Fig. 3.**
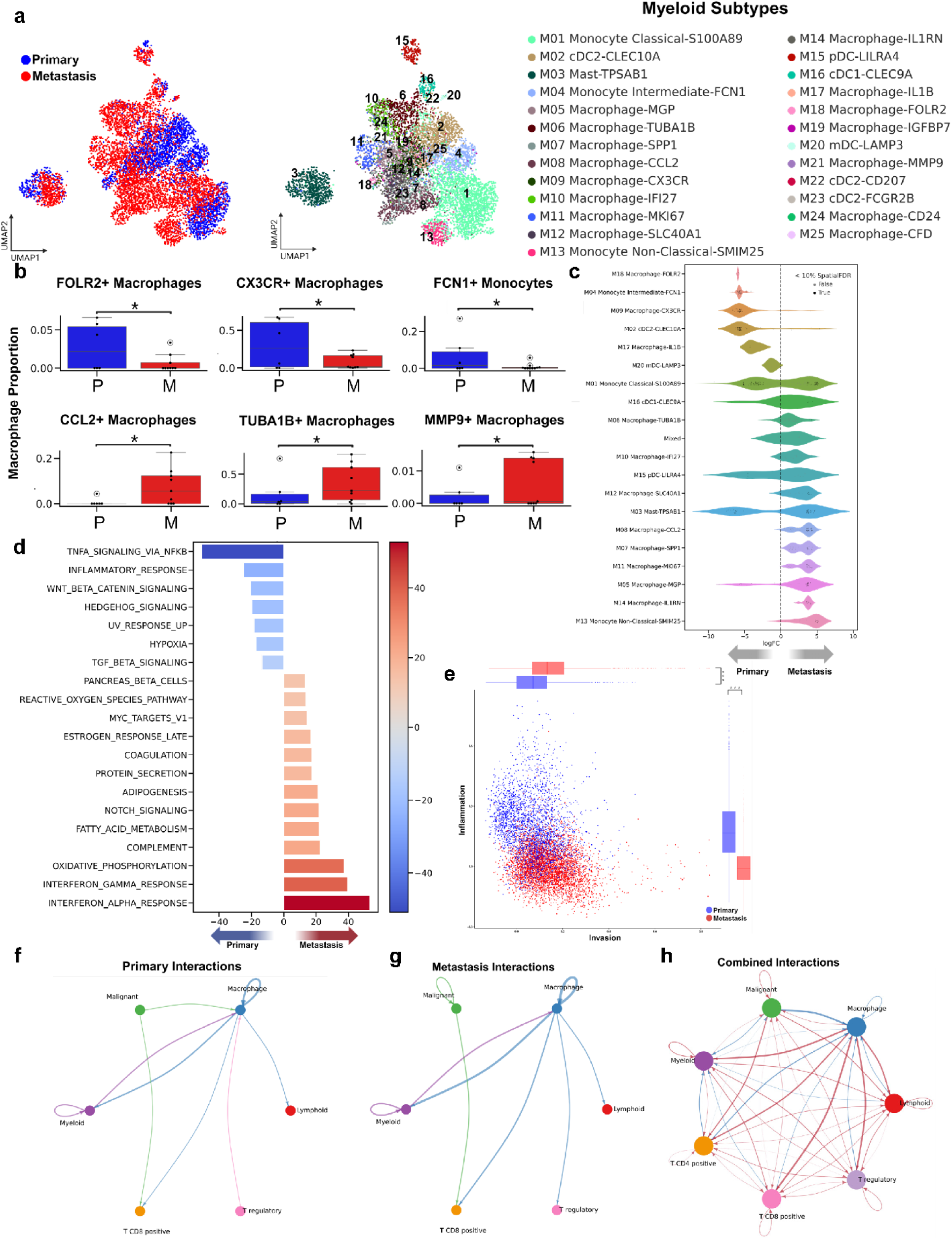
Distinct Myeloid Cell Populations and Their Roles in Primary and Metastatic Breast Cancer: Insights into Macrophage Polarization and Tumor Microenvironment Interactions. **a.** UMAP visualizations of 7651 myeloid cells. Cells are colored by tissue class of origin (left) and grouped by one of 25 identified sub-types (right), including 6 Dendritic, 3 Monocytic, 1 Mast, and 15 Macrophage lineages. **b.** Bar plots showing the proportions of cells from each tissue class of origin (P=Primary, M=Metastatic) for macrophage subtypes with a significant difference in proportion. (permutation test, p<0.05, log2FC > 2) **c.** The beeswarm plot illustrates the differential abundance analysis of myeloid cell subtypes, revealing distinct variations in cell abundance across neighborhoods, stratified by tissue classes of origin in primary and metastatic breast cancer. **d.** Differential Hallmark gene signature pathway activity scores within malignant cells across metastatic status. **e.** Cellular states plot showing CANCERSEA gene signature activity for Inflammation versus Invasion in myeloid cell populations. **f.** Circle plot displaying the top 15% of primary tumor microenvironment cell-cell interactions. Arrow colors indicate sender and thickness represents interaction strength. **g.** Circle plot displaying the top 15% of metastatic tumor microenvironment cell-cell interactions. Arrow colors indicate sender and thickness represents interaction strength. **h.** Comparative circle plot visualizing differential cell-cell communication between primary and metastatic tumor microenvironments, where red edges indicate increased interactions in metastatic tumors and blue edges denote increased interactions in primary tumors

Importantly, elevated expression of CXCL9 and CX3CR and also elevated FOLR2+ macrophages in tumors has been correlated with a better prognosis in breast cancer patients^16,18^. Importantly, CX3CR-expressing macrophages play a pivotal role in immune activation and have emerged as a promising target for cancer therapies due to their regulatory role in immune responses^58,59^. FCN1-positive monocytes have been associated with increased inflammatory function in several other cancer types, indicating a potential role for FCN1-expressing monocytes in modulating immune responses in cancer contexts^60^.

In the metastatic tumors, our study predominantly found an increase in macrophages expressing CCL2, MGP, SPP1, and MMP9. CCL2-expressing macrophages can potentially contribute to facilitating tumor cell invasion and metastasis^20,61–63^. This is supported by the role of the CCL2/CCR2 axis in TAM development, the direct impact of CCR2 signaling on tumor cell survival/growth and invasion/metastasis, and the association between high CCL2 expression and poor prognosis in cancer patients^20^.

Macrophages expressing MGP have been found to contribute to upregulating pro-tumorigenic factors associated with promoting immunoresistance^64^. SPP1+ tumor- associated macrophages were associated with poor prognosis in various cancers, potentially by contributing to tumor invasion by degrading the basement membrane through MMP expression^60^. Additionally, MMP9 expressing TAMs have been found to significantly contribute to creating a favorable environment for cancer metastasis^65^.

These macrophages have been linked to promoting aggressiveness and poor prognosis in various types of cancer^60,65^.

Using pseudobulk differential gene expression analysis, we observed a distinct partitioning of marker enrichment across myeloid cell populations exemplified by a volcano plot representation of differentially expressed genes. Intriguingly, the top-ranking genes associated with metastasis were predominantly linked to the interferon (IFN) response, underscoring the potential role of this pathway in the metastatic cascade. Indeed, chronic IFN signaling has been associated with an immunosuppressive immune contexture^66^. Conversely, the genes most prominently expressed in primary tumors were correlated with the TNF-α/NF-kB signaling pathway, suggesting a differential regulatory landscape between the primary and metastatic cellular contexts (Supp. Fig. 4a).

To gain a better understanding of the biological characteristics of myeloid cells, we investigated hallmark pathway activities within these cells. Our analysis revealed a significant increase in TNF-α signaling through the NFkB pathway in myeloid cells from primary tumors (Fig. 3d). In contrast, myeloid cells from metastatic tumors displayed pathways associated with oxidative phosphorylation, as well as responses to IFN-alpha and gamma. This suggests a diverse functional landscape across different myeloid cell states, which was also confirmed at the single-cell level (Fig. 3d, Supp. Fig. 4b).

We also utilized the CancerSEA^67^ database to examine cancer-related pathway activity. Our findings revealed a notable increase in invasion and metastasis traits in metastatic breast cancer macrophages, which implies a more aggressive phenotype in these cells, potentially contributing to the dissemination of the disease. In contrast, primary breast cancer myeloid cells showed a general increase in inflammation characteristics, particularly in the macrophage subset. This might suggest an active immune response within the primary tumor microenvironment, which could potentially influence tumor progression and treatment outcomes (Fig. 3e, Supp. Fig. 4c).

To delineate functional mechanisms in the immune microenvironment, we utilized CellChat^68^ to identify alterations in signaling interactions and cell-cell communications within the TME. Using CellChat, we focused on the top 15% predominant cell-cell interactions in both the primary and metastatic groups (Fig. 3f, 3g).

When comparing primary and metastatic datasets, we noticed unique patterns of cell-cell communication. In the primary dataset, there was a greater frequency of signaling from malignant cells, Treg cells, CD4+ T cells, and CD8+ T cells to macrophages, suggesting a strong interaction between these cell types in the primary tumor context. However, in the metastatic dataset, we observed a change in this communication pattern with an increase in signaling from macrophages to Treg cells and CD8+ T cells. This reversal in signaling direction suggests a potential change in the role of macrophages within the metastatic TME, possibly affecting the behavior of Treg and CD8+ T cells. These observations emphasize the dynamic nature of cell-cell interactions within the TME and the importance of understanding these changes in relation to tumor progression and metastasis. Collectively, these observed differences in myeloid/macrophage interactions point to a gain in the immunosuppressive TME within metastatic sites (Fig 3h).

We observed an enrichment of ligands that could potentially mediate cellular interactions in both primary and metastatic cancers (Supp. Fig. 4d, 4e). We noted elevated pro-inflammatory activity interactions, specifically with markers such as CXCL1, CXCL2, and CXCL8 in primary breast cancer. In the case of metastatic breast cancer, our study revealed aberrant interactions involving markers such as FN1, COL1A1, COL1A2, COL6A1, and THY1. These markers are integral to the formation and regulation of the ECM, a critical component of the tumor microenvironment. Aberrant interactions involving these proteins could lead to alterations in the ECM, potentially promoting cancer progression and metastasis^69,70^ (Supp. Fig. 4d, 4e).

### Aberrant lymphoid response within metastatic breast cancer TME

To expand upon our prior findings, which highlighted the interactions between immune cells and the TME, we aimed to identify possible alterations in lymphoid subtypes during breast cancer metastasis. We conducted an unsupervised clustering of lymphoid cells, resulting in the identification of twelve T cell, three B cell, and two NK cell subclusters (Fig. 4a) that clearly distinguished the subtypes in both primary and metastatic tumors.

**Fig. 4.**
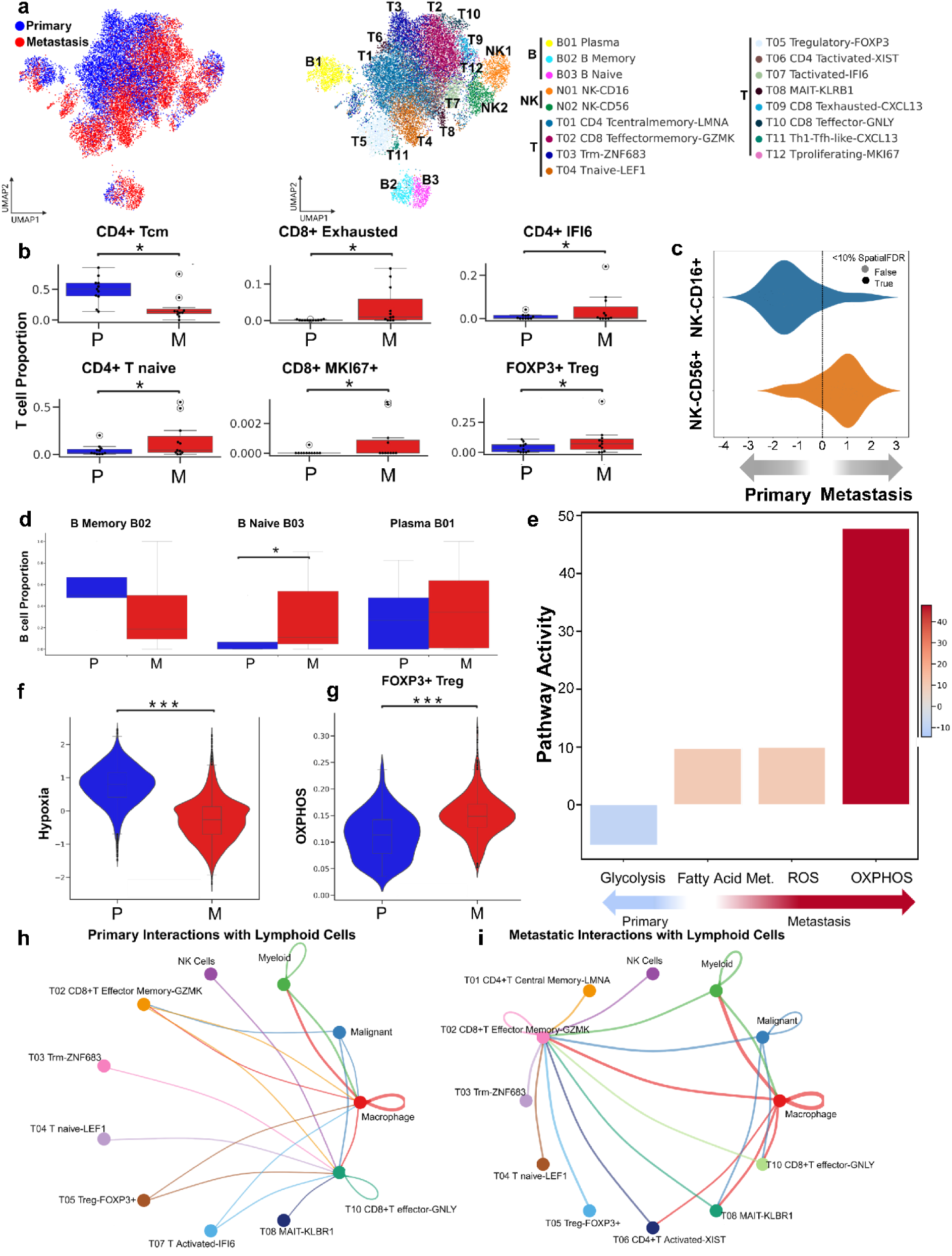
Lymphoid Cell Landscape and Functional Shifts in Primary vs. Metastatic Breast Cancer Tumors”. **a.** UMAP visualizations of 19104 lymphoid cells. Cells are colored by tissue class of origin on the left and grouped by one of 17 identified subtypes on the right, including 3 B cell subtypes, 2 NK cell subtypes, and 12 T cell subtypes. **b.** Bar plots showing the proportions of cells from each tissue class of origin (P=Primary, M=Metastatic) for T cell subtypes with a significant difference in proportion. (permutation test, p<0.05, log2FC > 2) **c.** The beeswarm plot illustrating the differential abundance analysis of NK cells, uncovering variations in cell abundance across neighborhoods, with a focus on specific NK cell subtypes, categorized by tissue classes of origin in primary and metastatic breast cancer. **d.** Bar plots showing the proportions of cells from each tissue class of origin (P=Primary, M=Metastatic) for B cell subtypes with a significant difference in proportion. (permutation test, p<0.05, log2FC > 2) **e.** Differential Hallmark gene signature pathway activity scores within T cells across metastatic status. **f.** Hypoxia score comparison between primary and metastatic breast cancer T cells. Significance indicated as (***, p < 0.001). **g.** OXPHOS score comparison between primary and metastatic breast cancer T cells. Significance indicated as (***, p < 0.001). **h.** Circle plot showing the top 10% of predominant cell-cell interactions in the primary tumor microenvironment, with arrow colors indicating the source (sender) of the interaction, and arrow thickness representing the strength of interaction. **i.** Circle plot showing the top 10% of predominant cell-cell interactions in the metastatic tumor microenvironment, with arrow colors indicating the source (sender) of the interaction, and arrow thickness representing the strength of interaction.

Although all T-cell subtypes were present in both primary and metastatic samples (Supp. Fig. 5a), the relative proportions of certain T-cell subtypes changed significantly from primary to metastatic tumors (Fig. 4b). Primary tumors generally displayed a higher proportion of pro-inflammatory T-cell subtypes, including CD4+ central memory T-cells that contribute to the rapid expansion of antigen-specific CD4+ T-cells and promote inflammation^71^. In contrast, metastatic samples exhibited mixed deregulation of T-cell subtypes, characterized by increases in CD8+ exhausted T-cells, CD4+ IFI6 T-cells, CD4+ Native T-cells, CD8+ MKI67+ T-cells, and FOXP3+ Tregs (Fig. 4b). The IFI6- expressing activated T cell (Tact) subtype is associated with disease progression post-chemotherapy in TNBC patients and correlates with a lower metastasis-free survival rate in breast cancer patients^21^. The upregulation of the IFI6 gene is linked to the control of mitochondrial ROS production, potentially contributing to unfavorable clinical outcomes in breast cancer patients^21^. These associations might also extend to the ER+ breast cancers in this study.

In our study, proliferative MKI67+ T cells were present exclusively in metastatic ER- positive patients. Studies in patients with triple-negative breast cancer (TNBC) with disease progression after chemotherapy have also shown proliferative MKI67+ T-cells (Tprf-MKI67)^21^. Additionally, MKI67 gene expression was significantly correlated with lymph node metastases, tumor invasion, and adverse survival outcomes in TNBC^21^.

In primary tumors, there is a higher proportion of CD16-positive NK cells, whereas in metastatic breast cancer, there is a higher proportion of CD56-positive NK cells (Fig. 4c). CD16-positive NK cells contribute to antibody-dependent cellular cytotoxicity (ADCC), a process where specialized immune cells, such as NK cells, recognize and kill cells coated with antibodies through interaction with CD16 receptors on their surface^72,73^. Additionally, our data revealed that primary tumors show significant enrichment in ADCC-related gene signatures compared to metastatic samples published previously^74^ (Supp. Fig. 5b).

Regarding B-cell subtypes, we found a decrease in memory B cells and an increase in naive B-cells within the metastatic samples (Fig. 4d). This observation is consistent with the active recruitment of B cells to the TME without prior priming against malignant cells.

### Lymphoid metabolic reprogramming results in an immunosuppressive metastatic TME

To further probe lymphoid subtypes, we performed pathway enrichment and also interaction analysis to decipher phenotypic changes. When considering all types of lymphoid cells, T cells in primary tumors show a preference for glycolytic signaling, while T cells in metastatic tumors are more enriched for fatty acid metabolism, reactive oxygen species production, and oxidative phosphorylation (Fig. 4e).

While our data shows higher hypoxic signaling in metastatic malignant cells, in primary samples, we rather observe this enrichment in the non-epithelial cell types. Indeed, we see a collective increase in hypoxic signaling, along with a decrease in oxidative phosphorylation in T cells from primary tumors compared to T cells from metastatic tumors (Fig. 4f, 4g). While low oxygen tension in T cells has been associated with inhibiting their function, recent studies also show that this characteristic can enhance aspects of the adaptive immune response^75^. Additionally, hypoxia during antigen recognition may differentially prime T cells, leading to improved antitumor activity^76,77^.

Network analysis revealed a dramatic shift in lymphoid cell-related signaling in metastatic tumors (Fig. 4h, 4i). Malignant and lymphoid interactions in primary tumors were dominated by an immunostimulatory regime, which largely signaled CD8+ T effector cells, while metastatic interactions were dominated by substantive signaling into CD8+ T effector memory (TEM) cells. The loss of the CD8+ cytotoxic population with a concomitant increase in CD8+ TEM may indicate a chronic immunosuppressive TME where CD8+ TEMs recognize malignant cells yet cannot effectively mediate tumor clearance^78,79^.

Our study observed varying levels of immune checkpoint inhibition across different cell types in both primary and metastatic breast cancers. Specifically, Tregs and Th1-like CXCL13+CD4+ T cells showed elevated expression of immune checkpoint transcripts. Also, metastatic lesions displayed a stronger immune checkpoint inhibition signal than primary tumors (Supp. Fig. 5c).

Th1-like CXCL13+ CD4+ T cells are known to interact with Tregs and are involved in downregulating genes associated with TCR signaling, similar to Tregs^80^. We also found that both cell types were more prevalent in metastatic samples (Supp. Fig. 5a). Our findings suggest that ER+ metastatic breast cancer patients may benefit from immune checkpoint inhibitor therapy, which could potentially enhance antitumor immune responses.

### Stromal cell remodeling occurs in the TME in metastatic sites

As key modulators of the stromal landscape of TMEs, we then focused on the characterization of fibroblasts. Unsupervised clustering of stromal cells revealed changes between primary and metastatic samples across one endothelial and 10 CAF subtypes (Fig. 5a) with a loss of both antigen-presenting (apCAF) and inflammatory CAFs (iCAF) coupled with a gain of both matrix CAFs(mCAF) and pericytes (Fig. 5a, 5b, Supp. Fig. 6a) in metastatic tumors.

**Fig. 5.**
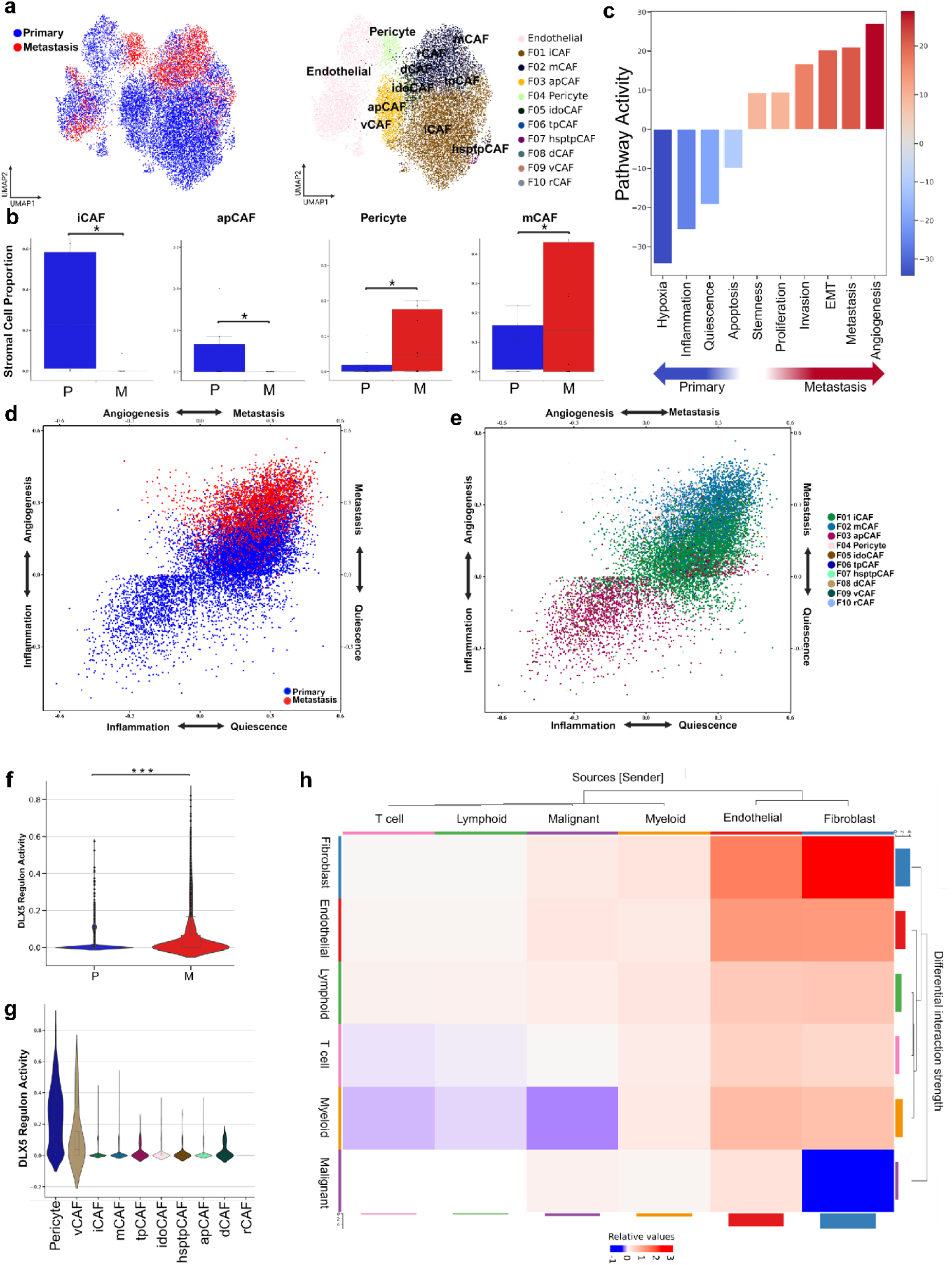
Stromal Cell Remodeling and Fibroblast Subtype Dynamics in Metastatic vs. Primary Breast Cancer TME. **a.** UMAP visualizations of 17339 stromal cells. Cells are colored by tissue class of origin on the left and grouped by one of 11 identified subtypes on the right, including 10 Fibroblast and an endothelial cell subtype. **b.** Bar plots showing the proportions of cells from each tissue class of origin (P=Primary, M=Metastatic) for Fibroblast cell subtypes with a significant difference in proportion. (permutation test, p<0.05, log2FC > 2) **c.** Differential CANCERSEA gene signature pathway activity scores within fibroblast cells across metastatic status. **d.** Cellular states plot to show CANCERSEA gene signature activity for Angiogenesis, Metastasis, Inflammation, and Quiescence across the metastatic status within fibroblast cells. **e.** Cellular states plot to show CANCERSEA gene signature activity for Angiogenesis, Metastasis, Inflammation, and Quiescence across the multiple fibroblast subtypes. **f.** DLX5 Regulon Activity score comparison between primary and metastatic breast cancer stromal cells. Significance indicated as (***, p < 0.001). **g.** DLX5 Regulon Activity score comparison between primary and metastatic breast cancer stromal cells. Significance indicated as (***, p < 0.001). **h.** Heatmap comparing the differential number of interactions and interaction strength between primary and metastatic tumor microenvironments. The top-colored bar plot indicates incoming signaling, while the right-colored bar plot represents outgoing signaling for each cell type. Red indicates increased signaling in metastatic tumors, whereas blue represents increased signaling in primary tumors. Bar height reflects the magnitude of change in interaction number or strength between the two conditions.

While CAFs are usually considered to have an immunosuppressive role^81^, antigen-presenting cancer-associated fibroblasts can enhance antigen presentation in the tumor microenvironment through MHC-II^82^. In primary breast cancer samples, there were higher levels of inflammatory CAFs, which were identified by high expression of cytokines such as CXCL12, CXCL14, and IL6^83^. These cytokines are indicators of inflammation and suggest an increased pathway involving PDGF, STAT3, KRAS signaling, and complement activation^83^.

In metastatic tumors, we observed an increase in matrix cancer-associated fibroblasts, which exhibited gene expression associated with invasion, such as matrix metalloproteinases (MMP), consistent with the alteration of the metastatic breast cancer TME^83^. Vascular pericytes are also found in a higher proportion in metastatic breast tumors and have previously been found to play a part in cancer invasion and metastasis through several pathways^84^.

As seen with myeloid and lymphoid lineages, stromal cells similarly exhibited higher levels of hypoxic signaling within the primary TME (Fig. 5c), supporting the notion that hypoxic stress shifts from non-tumor cells to tumor cells in the metastatic sites. Our findings indicate that hypoxic signaling varies between these sites, reflecting differing responses to extracellular oxygen levels rather than uniformity across cell types^85^.

Metastatic stromal cells displayed a notable increase in genes associated with angiogenesis and metastasis, indicating their potential involvement in promoting the growth and spread of tumors (Fig. 5c, 5d, 5e). In contrast, fibroblasts from primary TME showed an increase in inflammation and quiescence-related gene signatures.

Additionally, primary breast cancer fibroblasts displayed distinct cellular states at a cellular level, with subtypes including iCAFs within the primary TME predominantly exhibiting a quiescent state (Fig. 5d, 5e).

Next, we investigated the impact of transcription factors on fibroblasts. We observed high regulon activity of DLX5, a member of the MMP family^86^, especially in vascular cancer-associated fibroblasts (Pericytes and vascular CAFs) (Fig. 5f). Notably, we observed significantly higher DLX5 regulon activity in metastatic samples compared to primary tumors (Fig. 5g).

We evaluated communication between stromal cells and other cell types using the CellChat framework (Fig. 5h). Specifically, we found that myeloid cells exhibited heightened intercellular communication with fibroblasts in primary tumors, which aligns with our analysis of communication between malignant and tumor cells. Additionally, we discovered that fibroblasts established communication channels with other cell types and within themselves, particularly in the metastatic TME.

To gain a deeper understanding of the signaling pathways influencing cell-cell interactions in the TME, we conducted network analysis utilizing the MultiNicheNet^87^ approach. Our analysis revealed significant integrin and collagen-mediated interactions among stromal cells, as depicted in (Supp. Fig. 6b, 6c). Integrins are known to play a critical role in cell-cell and cell-extracellular matrix interactions, while collagen is a major component of the extracellular matrix^88^. The presence of these interactions underscores the importance of integrin and collagen-mediated signaling in the communication and behavior of stromal cells within the TME, particularly in metastatic tumors^89^.

## Discussion

As primary breast cancers progress and transition to metastatic breast cancer, an imbalance of TME signaling occurs that ultimately leads to poorer outcomes^90^. Unraveling the interactions within the TME is an evolving need, where tools, including single-cell sequencing, can reveal complex interactions between cell types. Several recent studies have used sc-RNAseq to reveal changes among cellular players in TME, including in breast and pancreatic cancers^12,91^ with the breast cancer study being restricted to lymph node metastasis. In this study, we analyzed the cellular composition and signaling pathways in primary breast cancers in comparison to tumors occurring in the most prevalent metastatic sites, including lymph nodes, liver, bone, adrenal gland, and subcutaneous (Fig. 1a). Our differential gene expression analysis did not identify substantial differences between tumor metastatic sites. This could be due to the small sample size analyzed, as well as the possibility that metastasis-specific gene expression patterns have a high degree of conservation across the different sites. With our analysis, we shed light on the core cellular and functional differences between primary and metastatic breast cancer niches.

While the main cell types were consistent between primary and metastatic samples, there were significant differences in the proportions of cellular subtypes within each group. The substantial cellular heterogeneity highlights the need for precise treatment approaches, with the differences between the subtypes potentially indicating therapeutic opportunities.

We observed pronounced transcriptional dynamics within malignant cells, indicating significant diversity in gene expression between primary and metastatic tumors. Our analysis of CNVs revealed substantial inter-patient differences in CNV profiles within both primary and metastatic tumor cell populations, further supporting the presence of intertumoral heterogeneity. We identified specific chromosomal regions that were pronounced in metastatic samples, encompassing genes associated with the progression and aggressiveness of breast cancer^25–32^ (Supp. Fig. 2b). Consistent with previous studies^92^, metastatic tumor cells exhibited a higher average number of clones compared to primary tumor sites.

Additionally, our study examined the classification of malignant cells using Integrative Clusters (IntClust) derived from the METABRIC study^52^. We observed an enrichment of more aggressive IntClust types in metastatic breast cancer, including IntClust1, 2, and 9. These IntClust types have been associated with late-recurring ER-positive genomic subgroups, genomic alterations contributing to aggressive behavior, and amplification of the MYC oncogene, respectively. This enrichment further supports the notion that metastatic breast cancer displays a more aggressive tumor behavior compared to primary breast cancer. Interestingly, single-cell analysis indicated that the primary and metastatic breast cancers contain cells from multiple different IntClust types, with the designated IntClust likely representing the dominant cell type in the tumors. This also suggests that some aspects of the IntClust may be due to cell state differences rather than determined by the mutation status of the cell.

While studying differences in activity of signaling pathways between the two disease states, we found elevated TNF-α via NFkB signaling pathway activities in primary breast cancer samples compared to metastatic samples. In contrast, metastatic samples showed higher JAK-STAT and IFN pathway activities. The activation of the TNF-α pathway in primary tumors may contribute to the recruitment of immune cells and the production of pro-inflammatory cytokines and chemokines^93^. Notably, the prominence of inflammatory signaling in primary tumors implies a potential role in initiating key processes conducive to tumor progression, including tumor cell invasion and migration^94^. Specifically, this aberrant signaling may serve as an early driver in the pathogenesis of primary tumors, setting the stage for subsequent metastatic dissemination^95^. The observed loss of NFkB signaling in metastatic malignant cells may contribute to the establishment of an immunosuppressive microenvironment permissive for growth in the metastatic niche^96^.

Our findings highlight the heterogeneity of myeloid cells, particularly macrophages, in the TME and point to their role in tumor progression and metastasis. In primary tumors, we observed that the enrichments of specific macrophage and monocyte subtypes are linked to better prognosis in cancer. While FOLR2+ macrophages were found to communicate more effectively with CD8+ T cells^16^, CXCR3+ macrophages, on the other hand, play a pivotal role in immune activation and have emerged as a promising target in cancer therapies. The diminished expression of CXCR3 in macrophages in metastatic breast cancer may be linked to changes in the immune microenvironment that support tumor survival in the metastatic niche^59^. FCN1+ monocytes have also been associated with increased inflammatory function in other cancer types, suggesting their potential role in modulating immune responses in cancer contexts^60^.

Our study further revealed the presence of macrophages expressing CCL2, MGP, SPP1, and MMP9 in metastatic tumors. As described above, these TAMs have been associated with aspects of tumor metastasis and may prepare metastatic sites for malignant cell growth.

Our examination of marker enrichment and pathway activities within myeloid cells revealed clear differences between primary and metastatic tumors. Primary tumors showed heightened TNF-α signaling through the NFkB pathway, indicating an active immune response within the primary tumor microenvironment. On the other hand, metastatic myeloid cells exhibited pathways related to oxidative phosphorylation and responses to interferon-alpha and gamma. Acute Type I IFN signaling response is considered a key driver for inflammation^97^. Studies suggest that Type I IFN has a dual role in cancer and chronic inflammation. The Type I IFN response in the Tumor Microenvironment may promote pro-tumorigenic TAM infiltration in metastatic lesions^98^. However, chronic interferon stimulation has been proposed to be immunosuppressive^66^ and may contribute to the immunosuppressive microenvironment observed in metastatic ER+ breast cancers in our study. This diverse functional landscape across different myeloid cell states highlights the dynamic nature of the TME and its potential influence on tumor progression and treatment outcomes.

Furthermore, our investigation of cell-cell interactions within the TME has uncovered significant changes in cellular interactions in metastatic tumors. In the case of metastatic breast cancer, cell-cell interactions predominantly revolved around the formation and regulation of the ECM, a pivotal component of the TME. This suggests a significant shift in the role of macrophages within the metastatic TME, potentially influencing the behavior of other immune cells. These findings underscore the vital significance of understanding the dynamic nature of cell-cell interactions and their impact on tumor progression and metastasis.

Within the lymphoid context, primary tumors exhibited a higher proportion of pro-inflammatory T-cell subtypes, while metastatic samples displayed mixed deregulation of T-cell subtypes, including increases in exhausted T cells, IFI6 T cells, naive T cells, MKI67+ T cells, and Tregs suggesting a shift towards a chronic immunosuppressive tumor microenvironment.

Our study highlights the potential prognostic use of specific T cell subtypes in breast cancer. The presence of IFI6-expressing activated T cells was associated with disease progression post-chemotherapy and lower metastasis-free survival rates^21^. Additionally, the presence of proliferative MKI67+ T cells was exclusive to TNBC patients experiencing disease progression after chemotherapy, and their gene expression correlated with lymph node metastases, tumor invasion, and adverse survival outcomes. Our study suggests that similar processes may be involved in ER+ tumors.

In terms of NK cells, CD16-positive NK cells, which play a role in ADCC, were more prevalent in primary breast cancer. However, in metastatic breast cancer, there was a higher proportion of CD56-positive NK cells. This shift may suggest a loss of NK cell ADCC ability during the metastatic transition, consistent with previous studies^72,73^.

We observed a decrease in memory B cells and an increase in naive B cells within the metastatic samples. This shift suggests active recruitment of naive B cells to the metastatic tumor microenvironment without prior priming against malignant cells. These findings provide further evidence of the complex interactions between lymphoid cells and the tumor microenvironment during breast cancer metastasis.

The identification of specific T cell subtypes associated with disease progression and prognosis, as well as the dynamic changes in NK cell and B cell subtypes, provide valuable insights for the development of targeted therapies and immunotherapies in metastatic breast cancer. Further studies are warranted to elucidate the underlying mechanisms driving these alterations in lymphoid subtypes and their functional implications in the tumor microenvironment. We also investigated the metabolic reprogramming of lymphoid cells. We found that T cells in primary tumors showed a preference for glycolytic signaling, while T cells in metastatic tumors were more enriched for fatty acid metabolism, reactive oxygen species production, and oxidative phosphorylation. This indicates a shift in metabolic pathways during breast cancer metastasis. While low oxygen tensions in the T cell microenvironment have been associated with inhibiting their function, recent studies suggest that this characteristic can enhance aspects of the adaptive immune response and improve antitumor activity. Network analysis demonstrated a dramatic shift in lymphoid cell-related signaling upon metastasis. Primary breast cancer lymphoid interactions were dominated by an immunostimulatory regime, primarily signaling CD8+ T effector cells. In contrast, metastatic interactions were dominated by substantive signaling into CD8+ T effector memory cells. This suggests a chronic immunosuppressive TME in metastatic breast cancer, where CD8+ TEMs recognize malignant cells but cannot effectively mediate tumor clearance.

Finally, our study revealed varying levels of immune checkpoint inhibition across different cell types in both primary and metastatic breast cancer. Tregs and Th1-like CXCL13+CD4+ T cells showed elevated expression of immune checkpoint mediators. Metastatic lesions displayed a stronger immune checkpoint inhibition signal than primary tumors. These findings suggest that breast cancer patients with metastatic ER+ tumors may benefit from immune checkpoint inhibitor therapy, which could enhance antitumor immune responses^99^.

It has been established that endothelial and fibroblast cells play crucial roles in the tumor microenvironment. In the context of the TME, fibroblasts, specifically cancer-associated fibroblasts, have been recognized as key modulators of the stromal landscape. They interact with tumor cells and other stromal components, influencing tumor progression, immune response, and therapy resistance. Building upon this knowledge, our study delved deeper into the characterization of fibroblasts within the TME, particularly during metastasis. We observed a loss of apCAF and iCAF, coupled with a gain of mCAF and pericytes in metastatic breast cancer. While CAFs are generally considered to have an immunosuppressive role, apCAFs can enhance antigen presentation in the TME^82^. Notably, we observed higher levels of inflammatory CAFs, identified by high expression of cytokines associated with inflammation in primary breast cancer samples, indicating their potential involvement in promoting an inflammatory environment conducive to tumor growth. A subset of iCAFs in the primary samples showed an increase in quiescence-related gene signatures. The identification of these quiescent iCAFs suggests that these fibroblasts may have specific functions related to cellular dormancy and inflammation regulation within the primary TME^100^. In metastatic breast cancer samples, we observed an increase in matrix CAFs, which exhibited gene expression patterns associated with invasion, such as MMPs. This observation aligns with previous studies highlighting the role of fibroblasts in facilitating tumor invasion and metastasis. Moreover, the increased presence of vascular pericytes in metastatic breast cancer further implicates their contribution to cancer progression through various pathways.

We observed elevated hypoxic signaling in stromal cells within the primary TME, consistent with the increased hypoxia seen in myeloid and lymphoid lineages. Notably, hypoxic signaling levels varied across different cell types, underscoring the critical influence of oxygen availability in shaping the dynamics of the TME. This variation highlights the potential role of cell type-specific responses to hypoxia in driving tumor progression and adaptation.

One of the key findings in our study was the high activity of the DLX5 regulon, particularly in vascular CAFs and pericytes. DLX5 is known to regulate the expression of MMPs, which are enzymes involved in degrading the extracellular matrix and facilitating tumor cell invasion. The upregulation of DLX5 in metastatic stromal cells indicates a potential mechanism by which these cells contribute to the growth and spread of tumors. Overall, the high DLX5 regulon activity observed in vascular CAFs and pericytes in metastatic samples suggests its potential as a therapeutic target^86^. The prominent presence of integrin and collagen-mediated interactions in the metastatic stroma highlights their importance in establishing metastasis (Supp. Fig. 6b, 6c).

In conclusion, our study provides valuable insights into the characteristics and interactions of stromal cells within the TME. The differential hypoxic signaling, gene expression profiles, cellular states, and intercellular communication observed among stromal cells contribute to our understanding of tumor progression and metastasis. These findings may have implications for the development of targeted therapies aimed at disrupting the tumor-stroma interactions in cancer treatment.

## Methods

### Sample collection and preparation

This study was approved by the Oregon Health & Science University (OHSU) Institutional Review Board (IRB). All biospecimens were collected and analyzed under the OHSU IRB-approved CEDAR (IRB #20750) or MMTERT observational study (Mitri 2018) (IRB #16113).

Participant eligibility was determined by the enrolling physician, and informed written consent was obtained from all subjects. All biopsy biospecimen used in this study were prospectively collected freshly for single-cell RNA sequencing library construction.

### Tissue dissociation

Tumor tissues were disaggregated by gentleMACS kits (Miltenyi Biotec, #130-095-929) following the manufacturer’s protocol. The lysate was resuspended and filtered through a 70-µm cell strainer (130-098-462; Miltenyi Biotec Germany). Cells were collected by centrifuging (300 × g for 7 min at 4 °C) and resuspended at 700–1200 cells/µl. Live cells were isolated by EasySep Dead Cell Removal (Annexin V) Kit (STEMCELL Technologies, #17899).

### Single-cell RNA sequencing library construction

Single-cell suspensions were processed according to the 10xGenomics scRNAseq sample preparation protocol (Chromium Next GEM Single Cell 3’ Kit v3.1, 10xGenomics). The entire mixed cell population was further analyzed without sorting or enrichment for specific cell subtypes. Cell suspensions were uploaded into the Chromium controller, capturing GEMs that encapsulated an estimated 5,000-10,000 single cells per channel. Libraries were constructed from the amplified cDNA, and sequencing was performed on the Illumina NovaSeq 6000 platform. All steps were performed according to the manufacturer’s standard protocol.

### Processing and quality control of scRNA-seq data

To ensure high-quality data, we implemented three quality control measures on the raw gene-cell-barcode matrix for each cell: the proportion of mitochondrial genes (≤20%), unique molecular identifiers (UMIs), and gene count (ranging from 400 to 100,000 and 200 to 10,000, respectively) using Scanpy^101^. Doublets were identified and removed using the Scrublet^102^ package for each sample. Normalization of total counts per cell was performed using the *normalize_total* function in the Scanpy^101^ package in Python, followed by log-normalization with the *log1p* function. Clustering was conducted using the Leiden algorithm at a resolution of 1, as provided by Scanpy^101^.

### Alignment and raw expression matrix construction

Raw sequencing data were aligned to GRCh38 genome reference using 10X software CellRanger (Version 6.1.2) with default parameters.

### RNA velocity analysis

For RNA velocity analysis, the spliced and unspliced reads were counted using the velocyto.py^103^ package (v0.17.17) from aligned bam files generated by CellRanger. A separate loom file was generated and used to process each sample further.

### Annotation

Marker genes for each cluster were identified using a t-test implemented in Scanpy^101^. These marker genes were then used to annotate each cluster using publicly available databases such as CellMarker^13^ and PanglaoDB^14^ To determine the cell identities.

These annotations were later refined using the CellTypist^15^ package and its annotate function. The model ’Immune_All_High.pkl’ was specified, and majority voting was enabled. Through this process, seven major cell type clusters were annotated: epithelial cells, natural killer cells, myeloid cells, T cells, B cells, fibroblasts, and endothelial cells. To distinguish malignant cells, we calculated and identified large-scale chromosomal copy number variation (CNV) by inferCNV^22^ and CaSpER^23^ tools for each sample based on transcriptomes. T cells and myeloid cells were considered reference cells; epithelial cells that had differing CNV patterns and exhibited higher CNV scores were annotated as malignant cells.

Datasets were merged across different samples using SCVI^104^ based integration, the top 4000 highly variable genes were used to train the VAE models, with each biopsy as a covariate key. After training the initial VAE model, the annotated cell types were used to build an extended model with scANVI^105^ for better integration.

After integration, NK cells, myeloid cells, T cells, B cells, and fibroblasts were further classified using the following publicly available single-cell RNA seq datasets: NK cells (GSE212890)^106^, T, myeloid, and B cells (GSE169426)^107^, and fibroblasts (GSE103322, GSE132465, GSE154778, and GSE212966)^83^. This label transfer was performed using the scArches^108^ algorithm, following the best practices described previously^109^.

### Proportion Analysis

A proportion analysis was conducted using the scProportionTest^110^ package to compare the fractions of cells within different cell populations. This involved performing a permutation test to calculate a p-value for each cluster and obtaining a confidence interval through bootstrapping.

### Constructing gene regulatory networks

We used pySCENIC^39^ to construct gene regulatory networks. This involved employing the GRNboost2 method for network inference. We used the *cisTarget* function with the Human motif database v10 to enrich gene signatures and pruned based on cis-regulatory cues using default settings. AUC scores were used to assess regulon enrichment across single cells, and the regulon specificity score was used to compute differential regulon activity.

### Copy number profile and subclone inference

We used the SCEVAN^37^ to determine clonal structures from inferred copy-number alteration profiles. The multiSampleComparisonClonalCN pipeline was employed for intratumoral comparison among multiple samples, with T cells and myeloid cells considered as the reference cells.

### Pathway analysis

To assess the pathway activities, we employed decoupler-py^111^ and retrieved the gene sets from the Molecular Signatures Database (MSigDB)^56^, PROGENy^55^ database, and CancerSEA^67^ databases.

For each single-cell, pathway activity was inferred using the *decouple* function with default parameters applied to gene sets that included weight information for each gene. In cases where gene sets did not have weight information, we employed the *aucell* function with default parameters, also within the decoupler-py^111^ package.

### Data visualization

We used essential Scanpy^101^ functions to generate UMAPs, box plots, heatmaps, dot plots, and violin plots. Proportional change was analyzed and visualized by using scProportionTest^110^ and Pertpy^112^.

For each condition under consideration, we generated pseudobulk profiles in accordance with the official vignette for pseudo-bulk functional analysis using the decoupler-py algorithm. The decoupler-py^111^ *plot_barplot* function was employed to illustrate the top absolute value activities for pathway activities in each condition. Additionally, we utilized the *plot_targets* function to compare primary and metastatic gene expression profiles, focusing on the target genes associated with each PROGENy^55^ pathway.

Visualization of the dimension reduction of cellular states for pathway activities at the cellular level was performed using the SCpubr^113^ *do_CellularStatesPlot*. Gene signatures sourced from the Molecular Signatures Database (MSigDB)^56^ were utilized to create these visual representations.

### Differential abundance testing

Graph-based differential abundance analysis was performed using the Milo^114^ framework in Pertpy^112^ to compare the cellular composition between primary and metastatic samples. The analysis followed the tutorial provided in the Pertpy package and utilized integrated datasets for each major cell type.

### Cell-cell communication analysis

We investigated cell-cell communication using the CellChat v2^68^ tool to infer, visualize, and analyze cell-cell communication networks from scRNA-seq data. The analysis was conducted following the recommended pipeline provided by the CellChat authors^68^.

Furthermore, we utilized the MultiNicheNet^87^ package to conduct cell interaction analysis, predicting ligand-receptor interactions between different cell types in both primary and metastatic tumor microenvironments. The analysis followed the guidelines outlined in the MultiNicheNet vignette. We visualized each group’s top twenty-five ligand-receptor interactions using the *make_circos_group_comparison* function.

### Survival Analysis

Kaplan-Meier survival analysis was performed using publicly available RNA-Seq datasets in the KM plotter^33^, focusing specifically on ER+ tumors among 2,575 patients. We assessed the collective high or low expression of the following genes: ARNT, BIRC3, EIF2AK1, EIF2AK2, FANCA, HOXC11, KIAA1549, MSH2, MSH6, and MYCN.

### Methodology for IntClust Classification

We utilized the Cluster Independent Annotation (CIA)^53^ tool to compute scores for each IntClust and to classify each cell accordingly. The top 200 differentially expressed genes (DEGs) for each IntClust, as identified in the METABRIC^52^ study, were utilized as input for the CIA tool. Specifically, we utilized the *CIA_classify* function to calculate scores based on the expression levels of the selected DEGs and assign each cell to the most probable IntClust.

## Data availability

Raw single-cell RNA sequencing data for this study are available in the NCBI BioProject database under accession number PRJNA1140267. Processed single-cell RNA sequencing data and single-cell RNA seq objects can be found as follows: https://doi.org/10.5281/zenodo.13743373

## Code availability

The authors declare that all scripts supporting the findings of this study are available from the corresponding author upon reasonable request.

## Author Information

F.O., T.Y.O., and A.O. contributed equally to this work. G.B.M. and H.M. conceived and supervised the project. F.O., T.Y.O., A.O., G.B.M., and H.M. designed the study. F.O. performed the computational bioinformatics analysis. F.O. and T.Y.O. wrote the original draft., with A.O., M.J.R., G.B.M., and H.M. providing editing contributions. A.O., T.Y.O., M.J., F.B., and X.L. were responsible for collecting clinical samples and conducting experiments. G.B.M and H.M provided the final review of the manuscript. G.B.M. and H.M. acquired the funding. All authors contributed to data generation and interpretation. All authors read and approved the final manuscript.

Author List: Furkan Ozmen, Tugba Y. Ozmen, Aysegul Ors, Mahnaz Janghorban, Matthew J. Rames, Xi Li, Fariba Behbod, Gordon B. Mills, Hisham Mohammed.

## Ethics Declarations

This study was approved by the institutional ethics review board of OHSU. Written informed consent was signed for all experiments involving patients.

## Supplementary Information

All supplementary materials are available upon publication. The supplementary materials include three spreadsheets, and one document as follows:

Supplementary Table 1: Clinical information related to samples in the study (Supplementary Table 1.xlsx)

Supplementary Table 2: Subclone information of malignant cells for individual samples (Supplementary Table 2.xlsx)

Supplementary Table 3: Proportions, counts, and pathway activity comparisons for various cell types across primary and metastatic breast cancer samples. This includes the proportion and count of major and minor cell types, differentially expressed markers, CNV and ITHGEX score distributions, IntClust subtype proportions, regulon activity scores, pathway activity comparisons (PROGENy, Hallmark, CancerSEA), and gene signatures used in the analysis (Supplementary Table 3.xlsx)

Supplementary Document 1: CNV alteration heatmaps for individual samples (Supplementary Document 1.pdf)

## Lead contact

Further information and requests for resources and reagents should be directed to and will be fulfilled by the Lead Contact, Furkan Ozmen.

## Supplementary Figures

**Supp Fig. 1.**
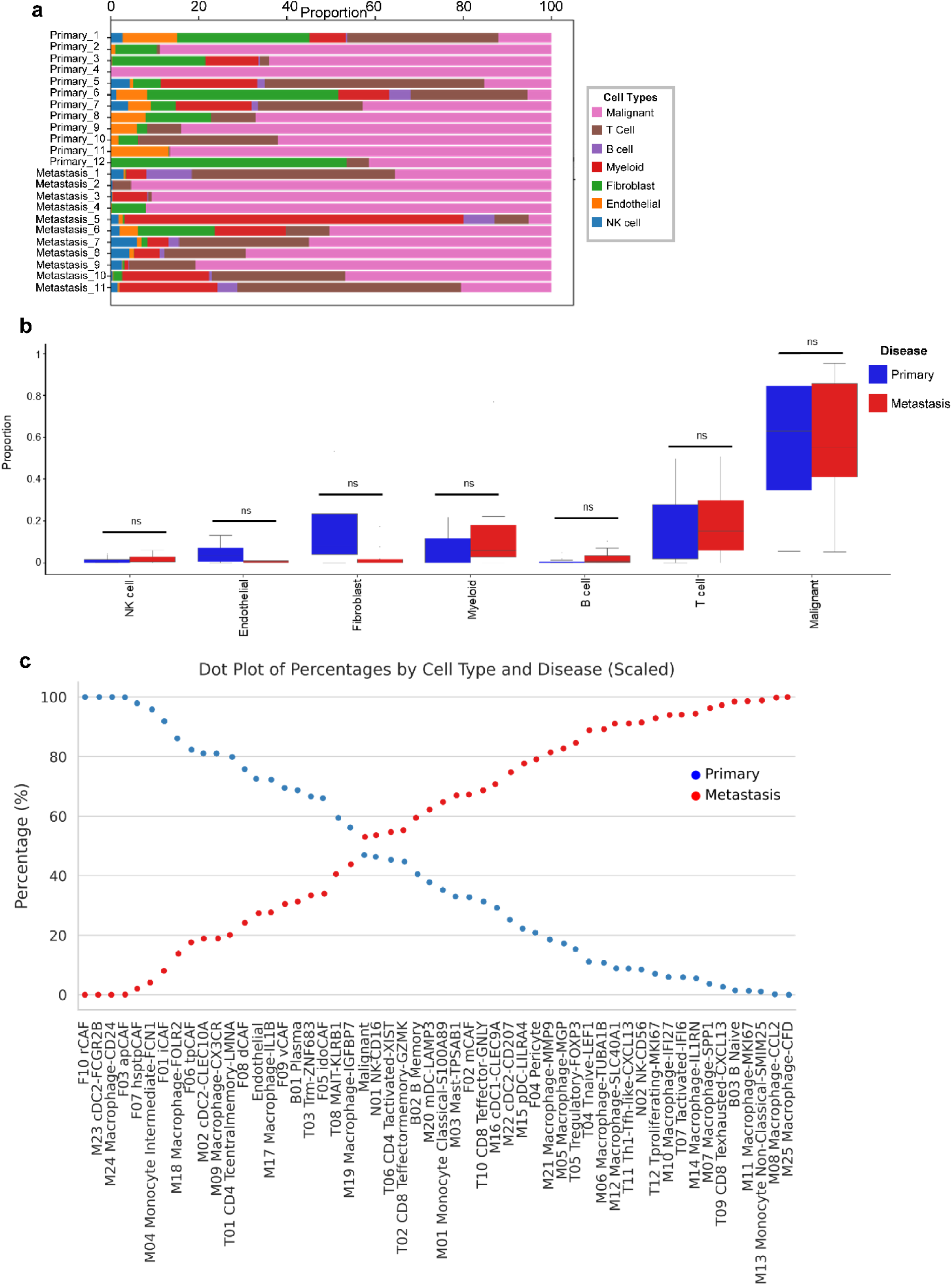
**a.** Stacked barplot showing the proportion of seven major cell types across individual samples. **b.** Boxplots illustrating changes in the proportions of seven major cell types between primary and metastatic breast cancer samples. (permutation test, p<0.05, log2FC > 2) **c.** Dot plot illustrating the percentage of each cell subtype from primary versus metastatic tumors, highlighting each cell subtype’s distribution and comparative abundance across the two conditions.

**Supp Fig. 2.**
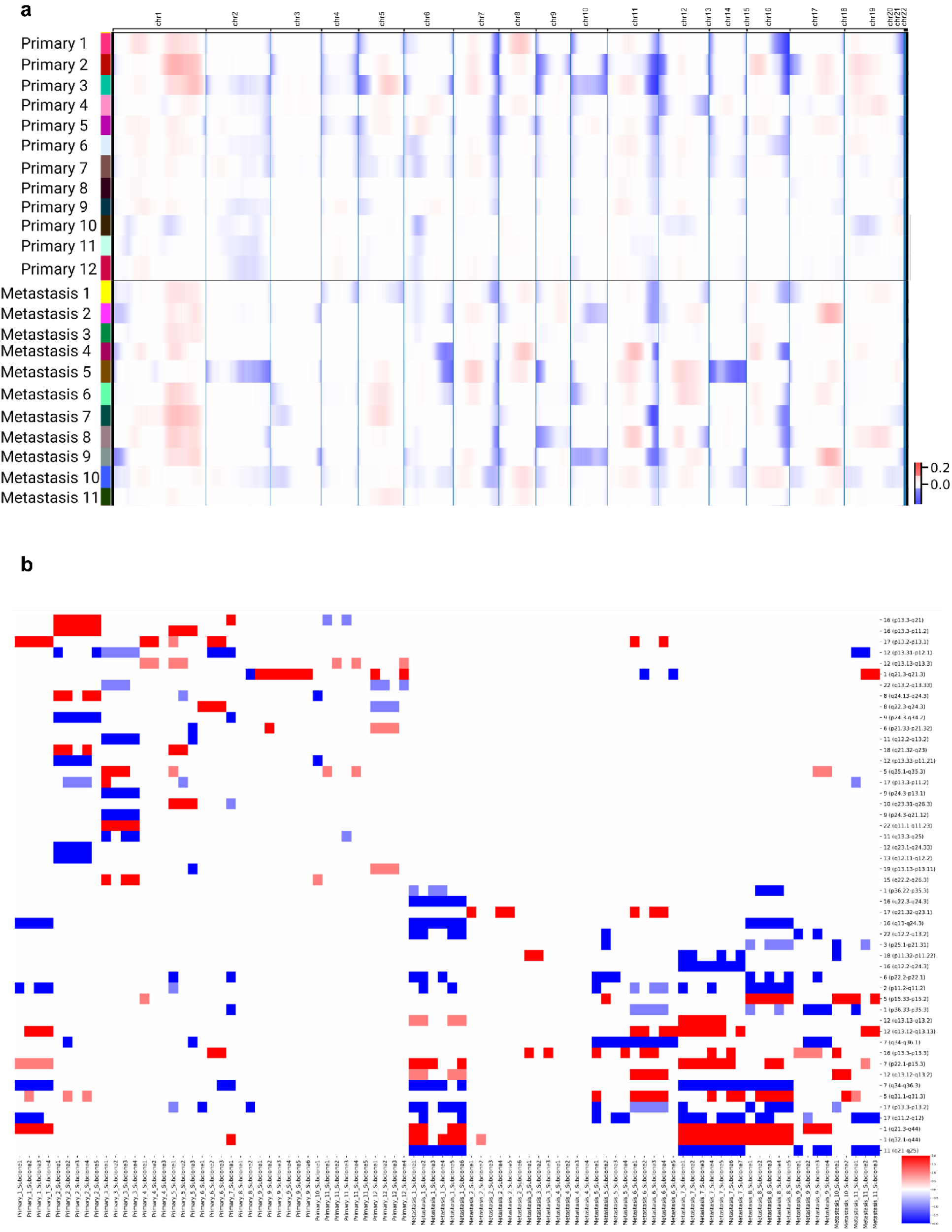
**a.** Heatmap showing CNV profiles for malignant cells in individual primary and metastatic tumor samples, with T cells as a reference. **b.** Heatmap depicting CNV profiles from all tumor subpopulations per patient, comparing the top 25 copy number alterations across chromosomal arms between primary and metastatic breast cancer subclones.

**Supp Fig. 3.**
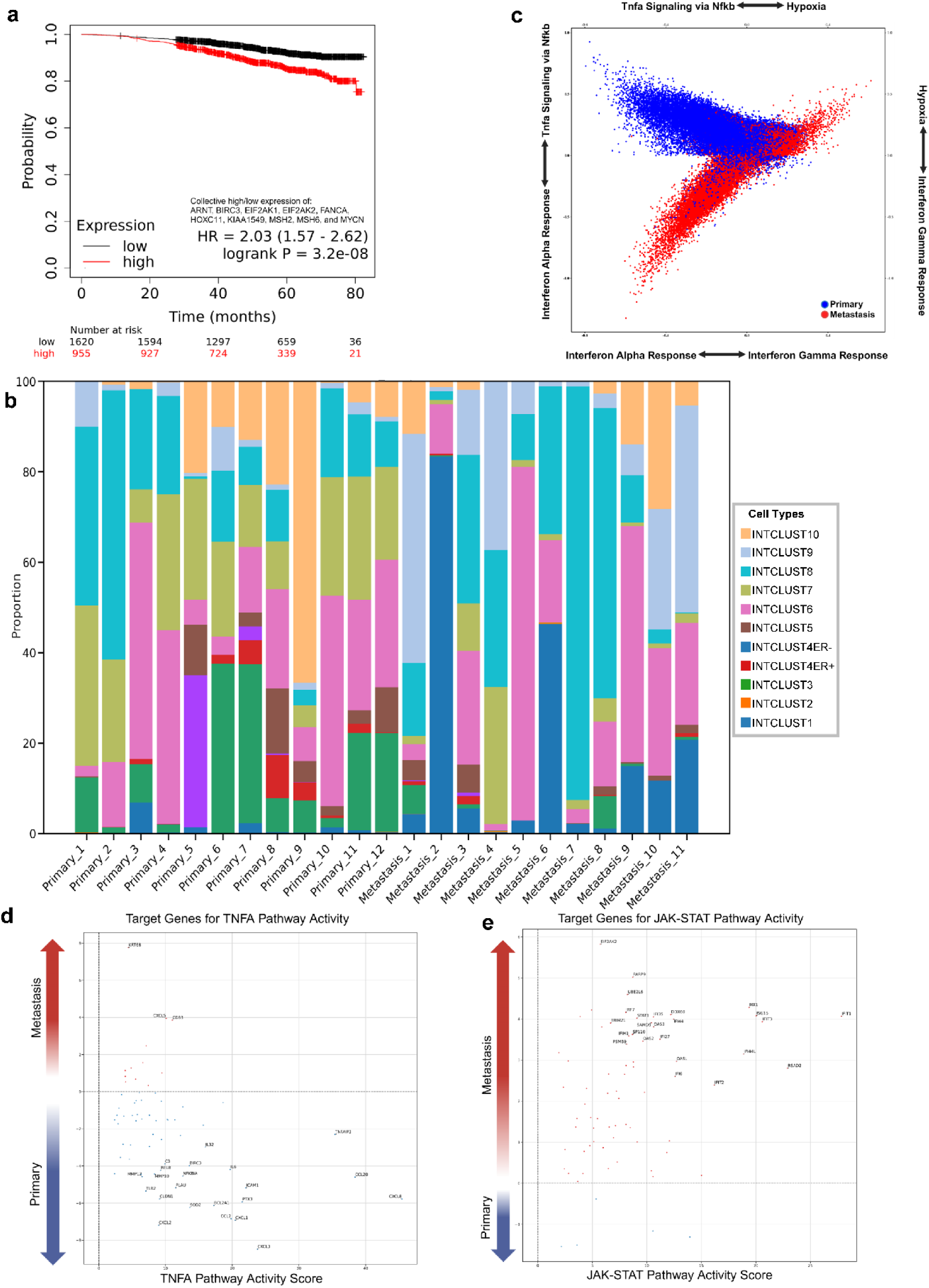
**a.** Kaplan-Meier survival curves for estrogen receptor-positive tumors among 2,575 patients, comparing survival outcomes associated with high versus low expression of the genes ARNT, BIRC3, EIF2AK1, EIF2AK2, FANCA, HOXC11, KIAA1549, MSH2, MSH6, and MYCN. **b.** Stacked bar plot showing the proportion of Integrative Clusters (IntClust) across individual samples, based on classification using the top 200 differentially expressed genes from the METABRIC study. **c.** Cellular states plot to show Hallmark gene signature activity for Hypoxia, Interferon Gamma Response, Interferon Alpha Response, and TNF-α Signaling via NFkB across the metastatic status within malignant cells. **d.** Plot of the top 25 target genes affected by TNF-α pathway activity, highlighting their weight and statistical significance in primary versus metastatic malignant cells based on PROGENY pathway activity. **e.** Plot of the top 25 target genes affected by JAK-STAT pathway activity, highlighting their weight and statistical significance in primary versus metastatic malignant cells based on PROGENY pathway activity.

**Supp Fig. 4.**
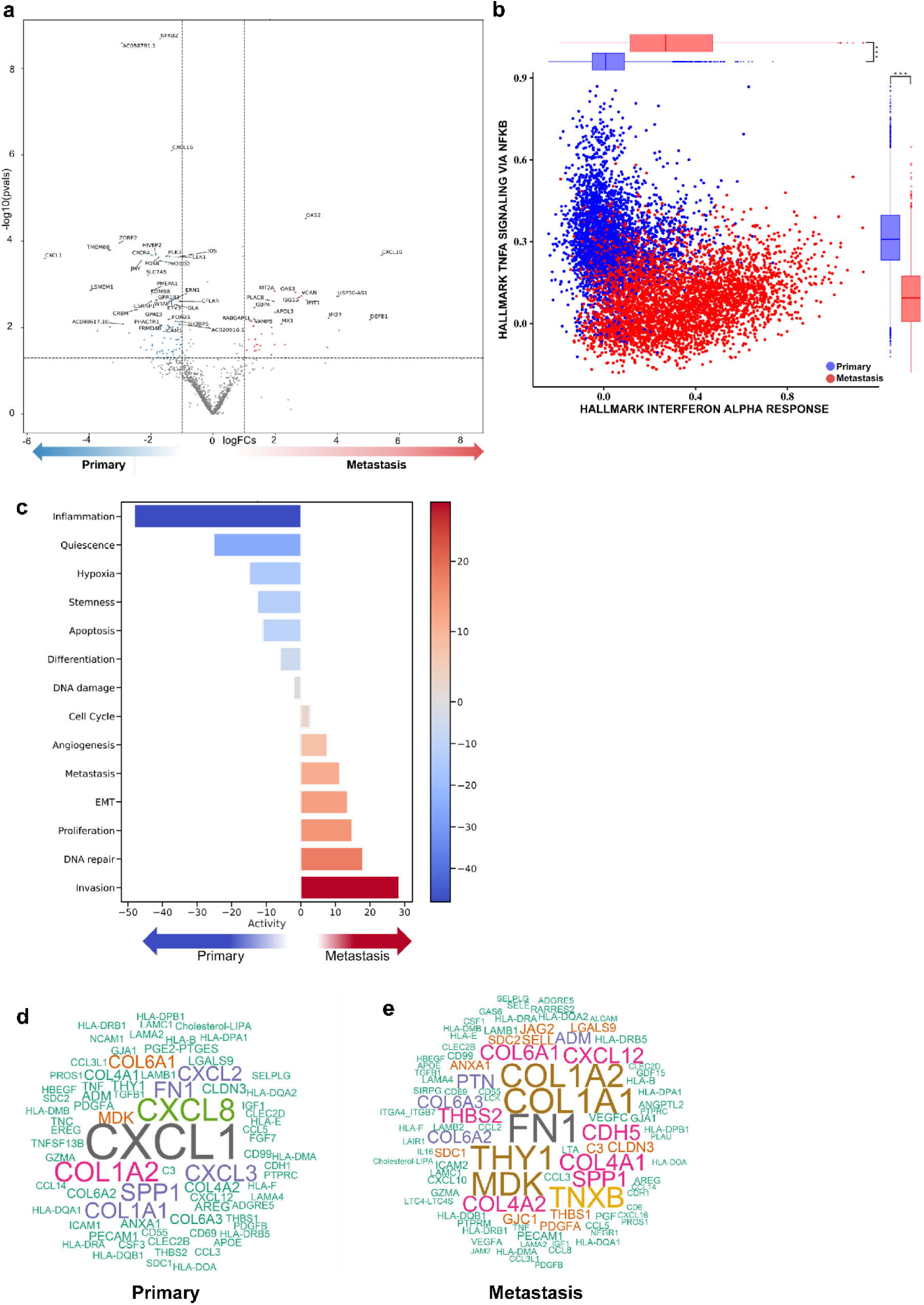
**a.** Volcano plot illustrating the differential expression of genes in myeloid cells when comparing primary versus metastatic tumors. **b.** Cellular states plot showing Hallmark gene signature activity for Interferon Alpha Response versus TNF-α Signaling via NFkB in myeloid cell populations. **c.** Differential CANCERSEA gene signature pathway activity scores within myeloid cells across metastatic status. **d.** Word cloud plot illustrating the enriched ligands in primary breast tumor microenvironment compared to metastatic cancer **e.** Word cloud plot illustrating the enriched ligands in metastatic breast tumor microenvironment compared to primary breast cancer

**Supp Fig. 5.**
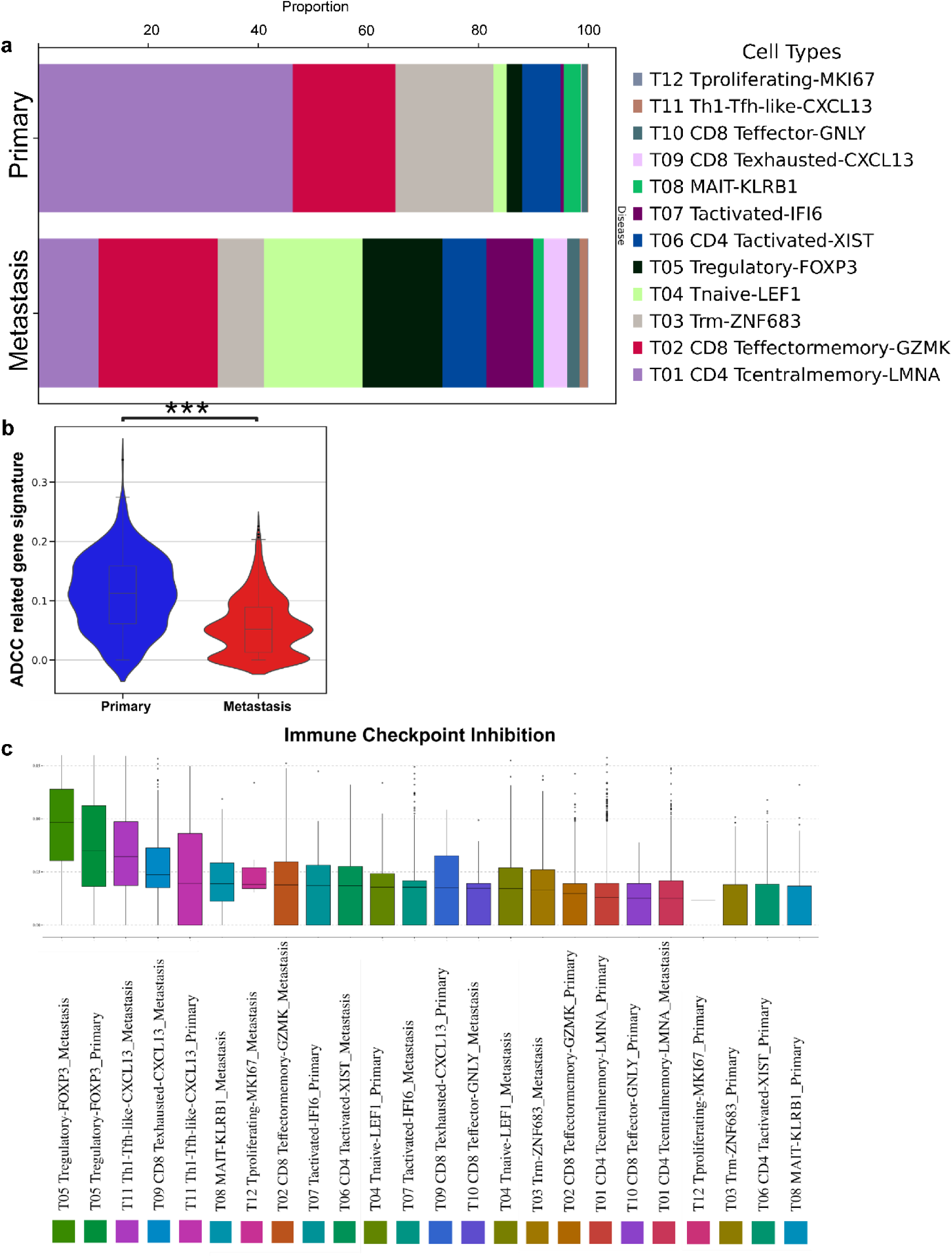
**a.** Stacked bar plot showing the proportion of T cell subtypes across the metastatic status. **b.** Antibody-dependent cellular cytotoxicity gene signature comparison between primary and metastatic breast cancer NK cells. Significance indicated as (***, p < 0.001). **c.** Immune checkpoint inhibition levels compared across different cell types in primary and metastatic breast cancers

**Supp Fig. 6.**
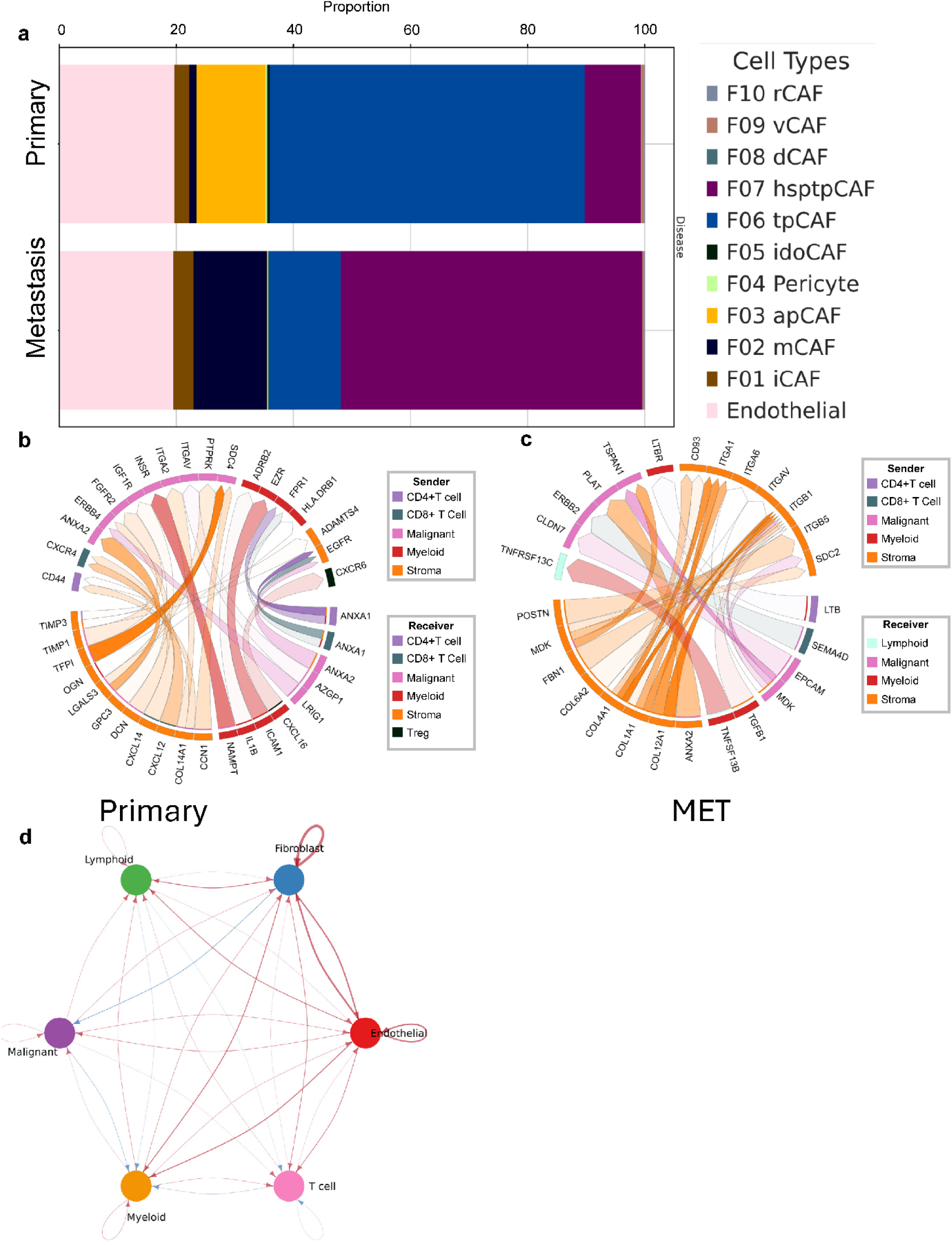
**a.** Stacked bar plot showing the proportion of stromal cell subtypes across the metastatic status. **b.** Visualization of the top 25 ligand-receptor interactions between different cell types in the primary tumor microenvironment, as analyzed using the MultiNicheNet. **c.** Visualization of the top 25 ligand-receptor interactions between different cell types in the metastatic tumor microenvironment, as analyzed using the MultiNicheNet. **d.** Comparative circle plot visualizing differential cell-cell communication between primary and metastatic tumor microenvironments, where red edges indicate increased interactions in metastatic tumors and blue edges denote increased interactions in primary tumors

