## Supplementary Document 1 for "Single-cell RNA sequencing reveals immunosuppressive pathways associated with metastatic breast cancer"

Primary 1

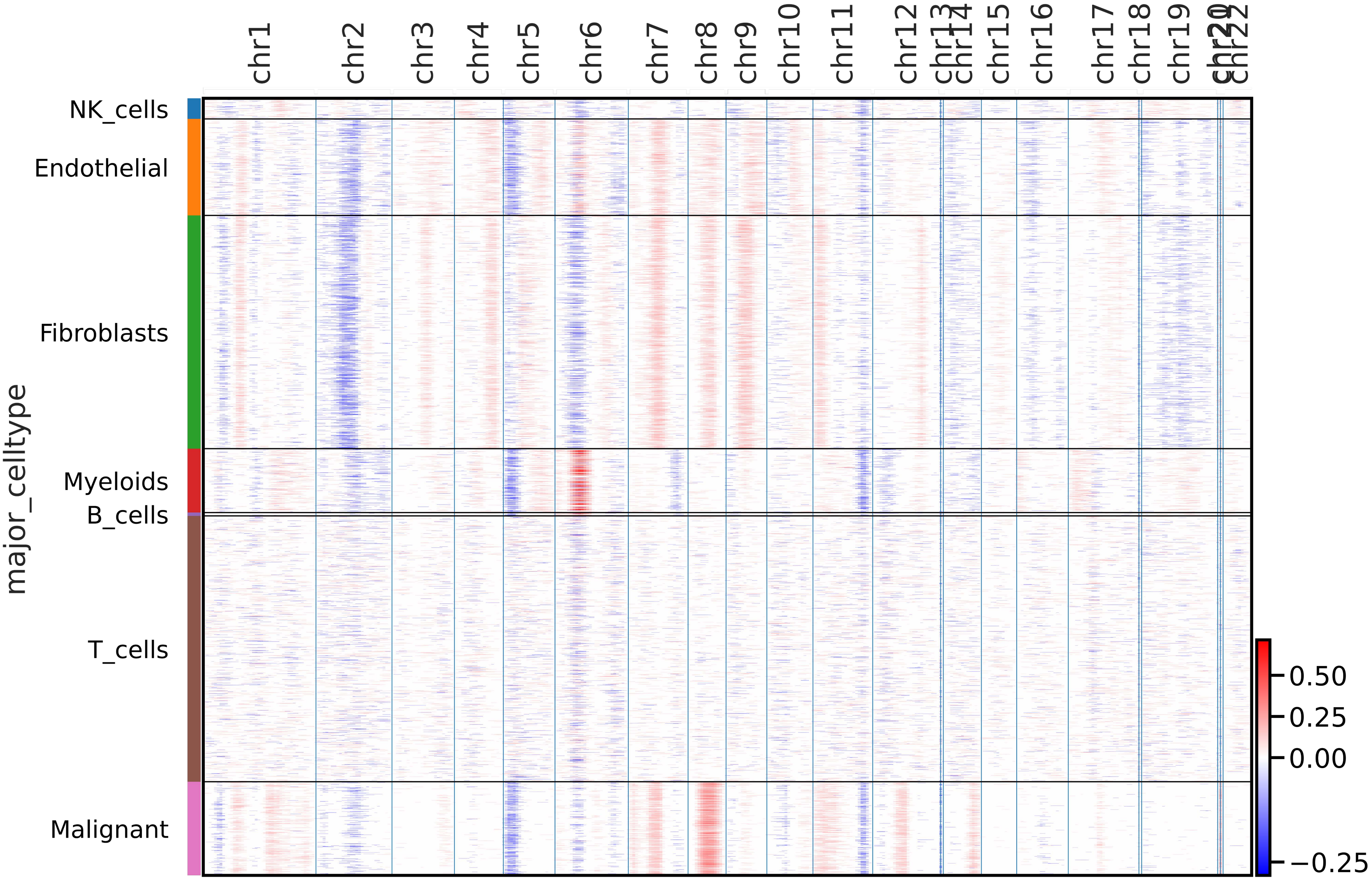

chr1
chr2
chr3
chr4
chr5
chr6
chr7
chr8
chr9
chr10
chr11
chr12
chr13
chr14
chr15
chr16
chr17
chr18
chr19
chr20
chr21
chr22
NK_cells
Endothelial
Fibroblasts
Myeloids
B_cells
T_cells
Malignant
major_celltype
0.50
0.25
0.00
−0.25

Primary 2

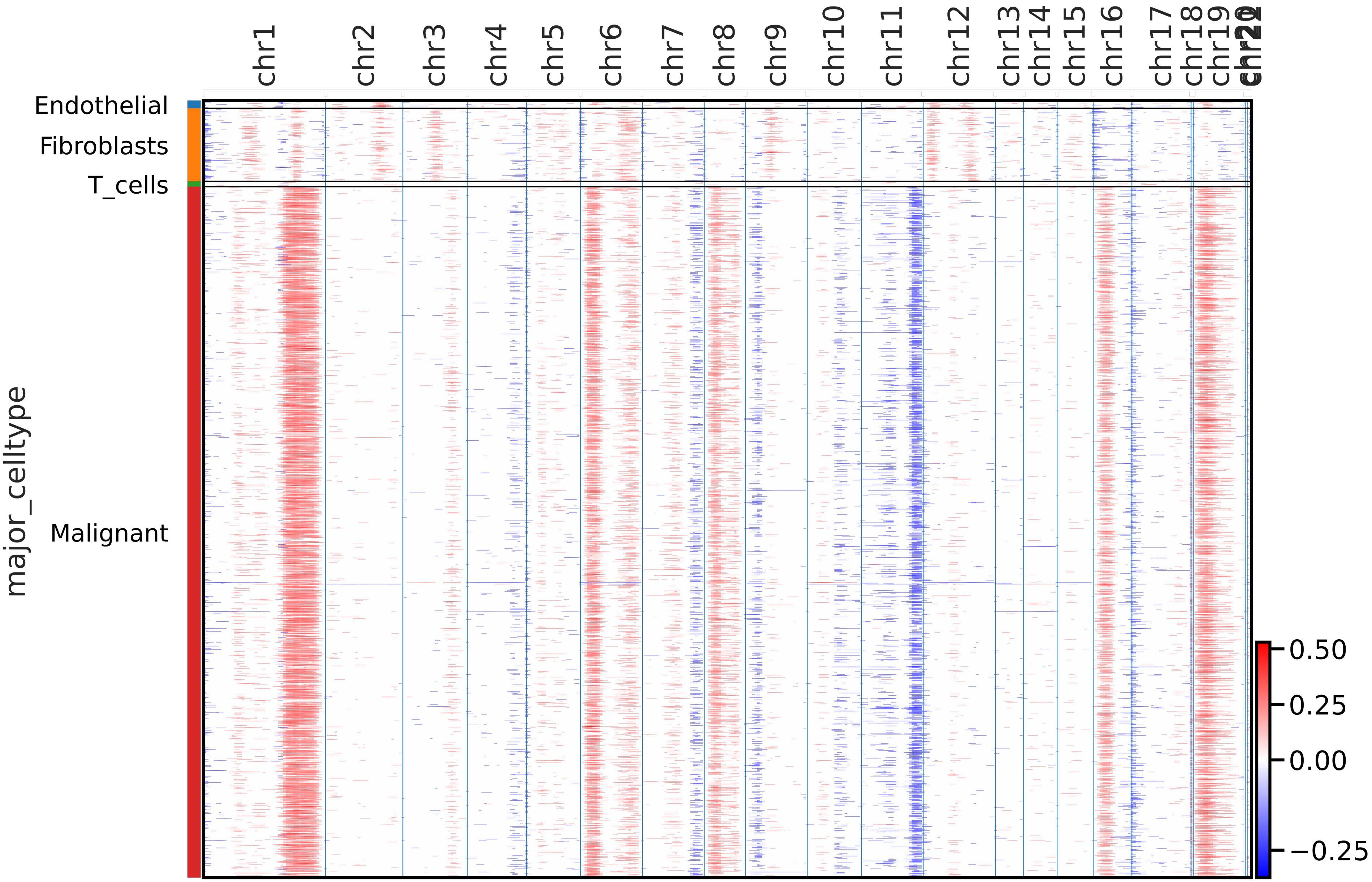
chr1
chr2
chr3
chr4
chr5
chr6
chr7
chr8
chr9
chr10
chr11
chr12
chr13
chr14
chr15
chr16
chr17
chr18
chr19
Endothelial
Fibroblasts
T_cells
Malignant
major_celltype
0.50
0.25
0.00
−0.25

Primary 3

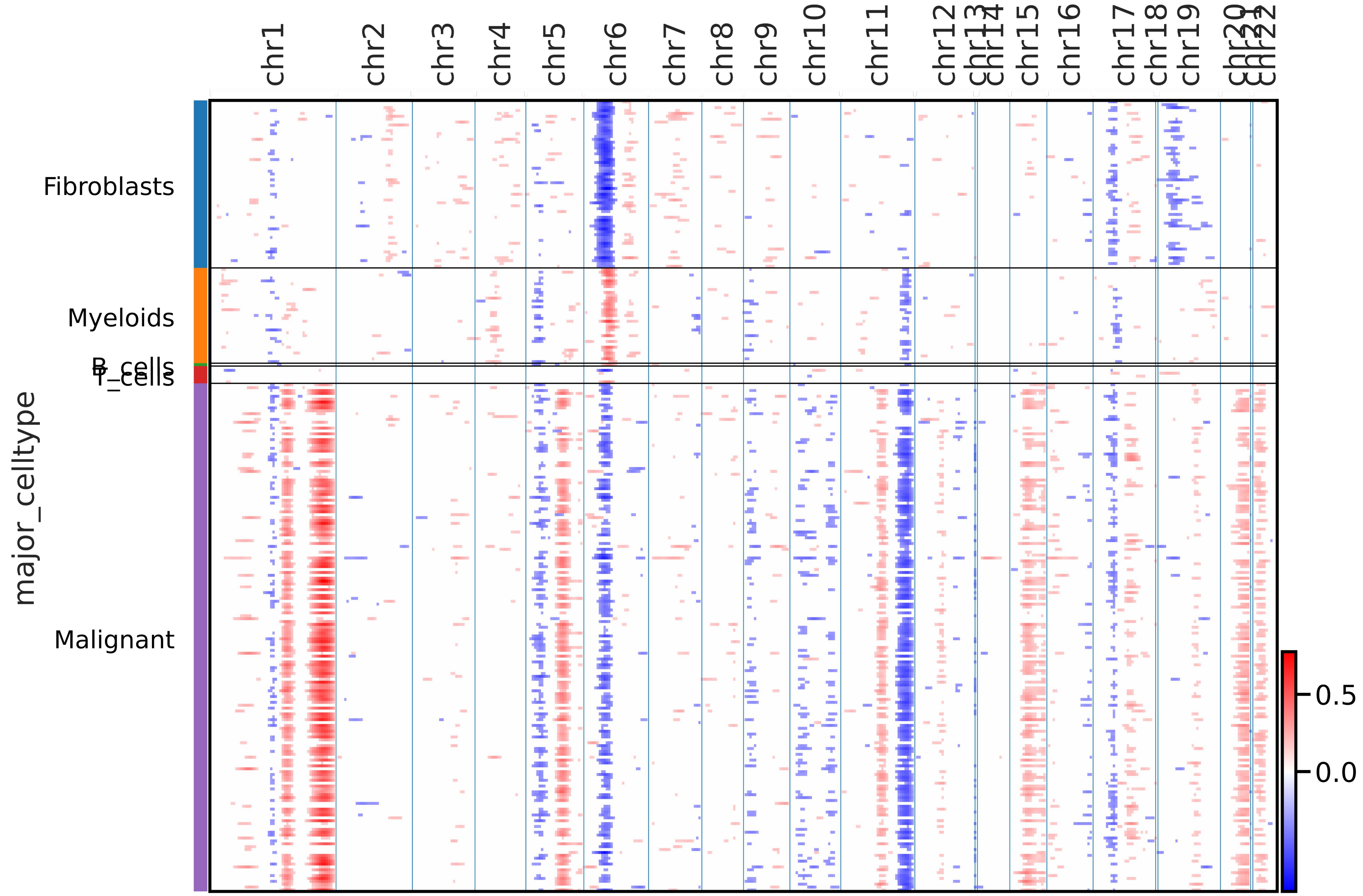

chr1
chr2
chr3
chr4
chr5
chr6
chr7
chr8
chr9
chr10
chr11
chr12
chr13
chr14
chr15
chr16
chr17
chr18
chr19
chr20
chr21
chr22
Fibroblasts
Myeloids
B_cells
T_cells
Malignant
major_celltype
0.5
0.0

Primary 4

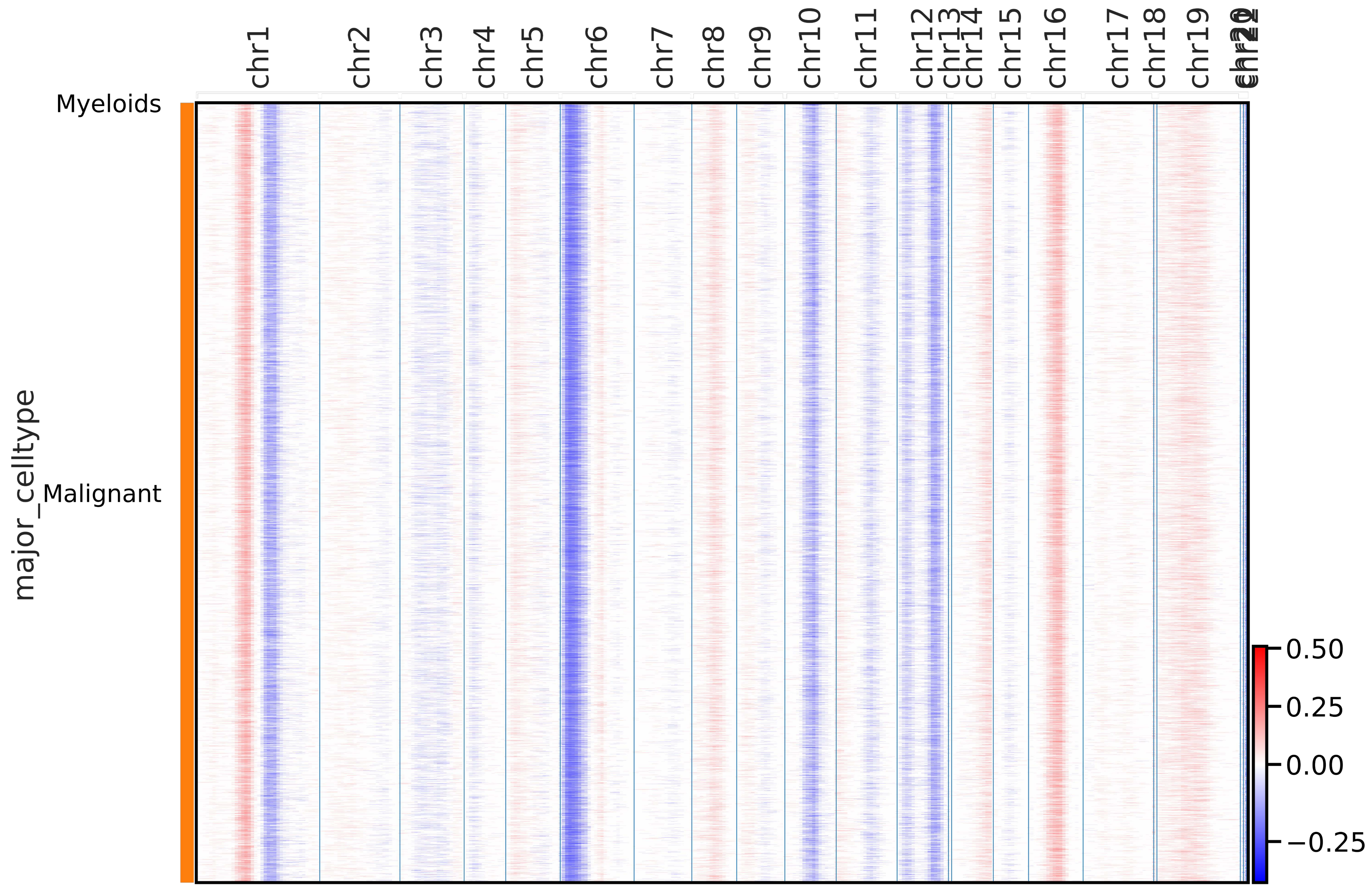

chr1
chr2
chr3
chr4
chr5
chr6
chr7
chr8
chr9
chr10
chr11
chr12
chr13
chr14
chr15
chr16
chr17
chr18
chr19
Myeloids
major_celltype
Malignant
0.50
0.25
0.00
−0.25

Primary 5

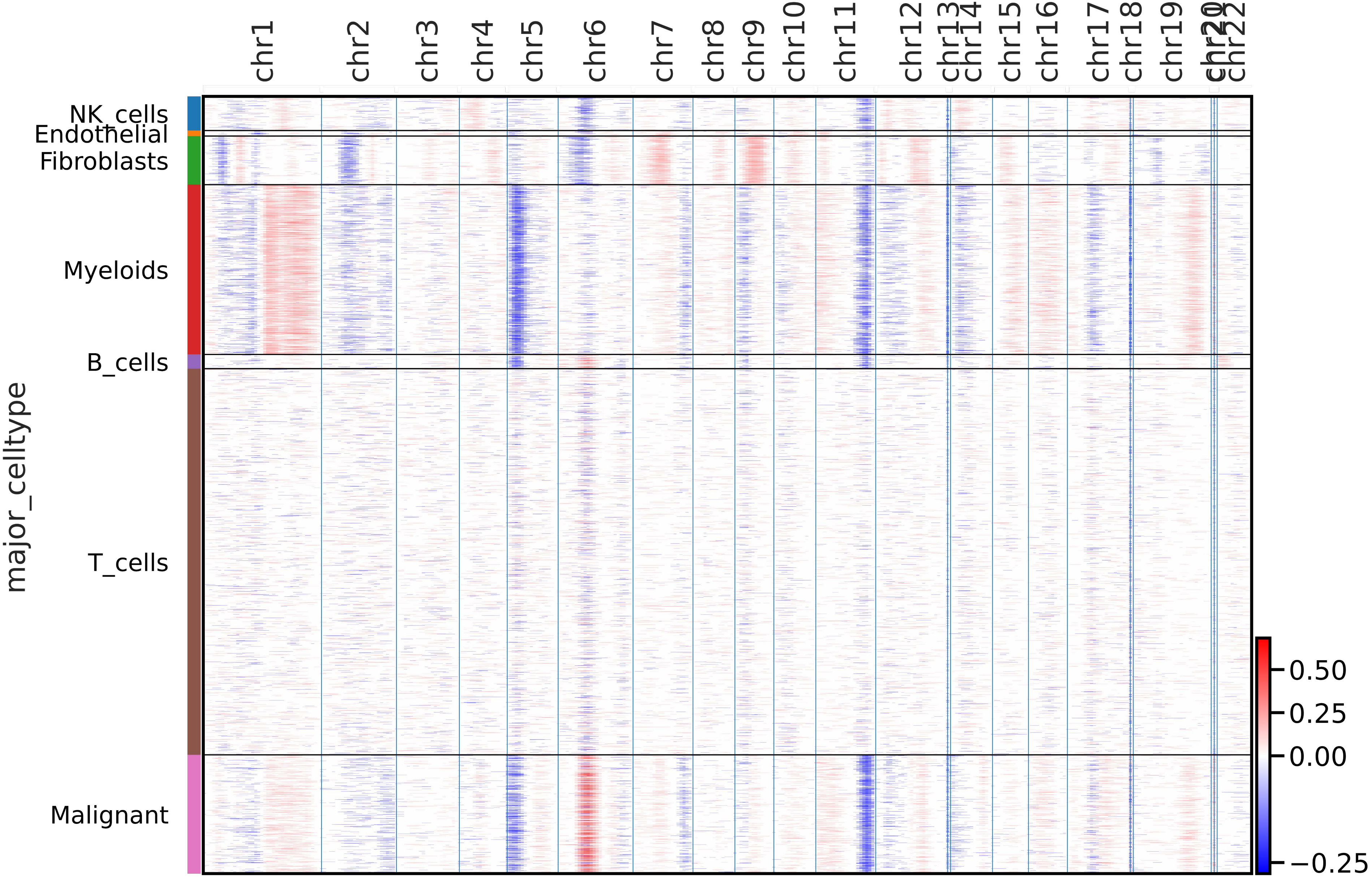

chr1
chr2
chr3
chr4
chr5
chr6
chr7
chr8
chr9
chr10
chr11
chr12
chr13
chr14
chr15
chr16
chr17
chr18
chr19
chr20
chr21
chr22
NK_cells
Endothelial
Fibroblasts
Myeloids
B_cells
T_cells
Malignant
major_celltype
0.50
0.25
0.00
−0.25

Primary 6

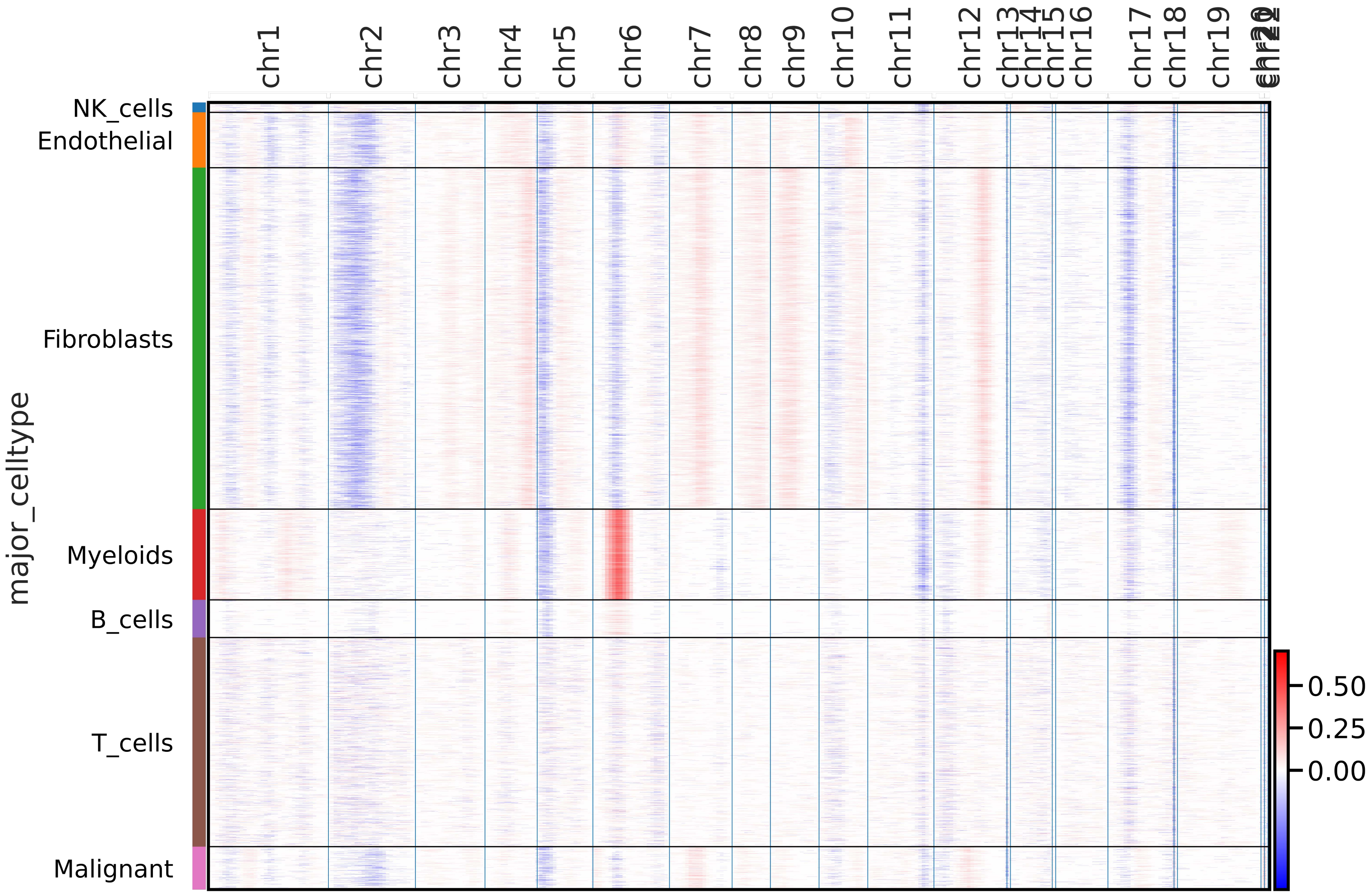

chr1
chr2
chr3
chr4
chr5
chr6
chr7
chr8
chr9
chr10
chr11
chr12
chr13
chr14
chr15
chr16
chr17
chr18
chr19
NK_cells
Endothelial
Fibroblasts
Myeloids
B_cells
T_cells
Malignant
major_celltype
0.50
0.25
0.00

Primary 7

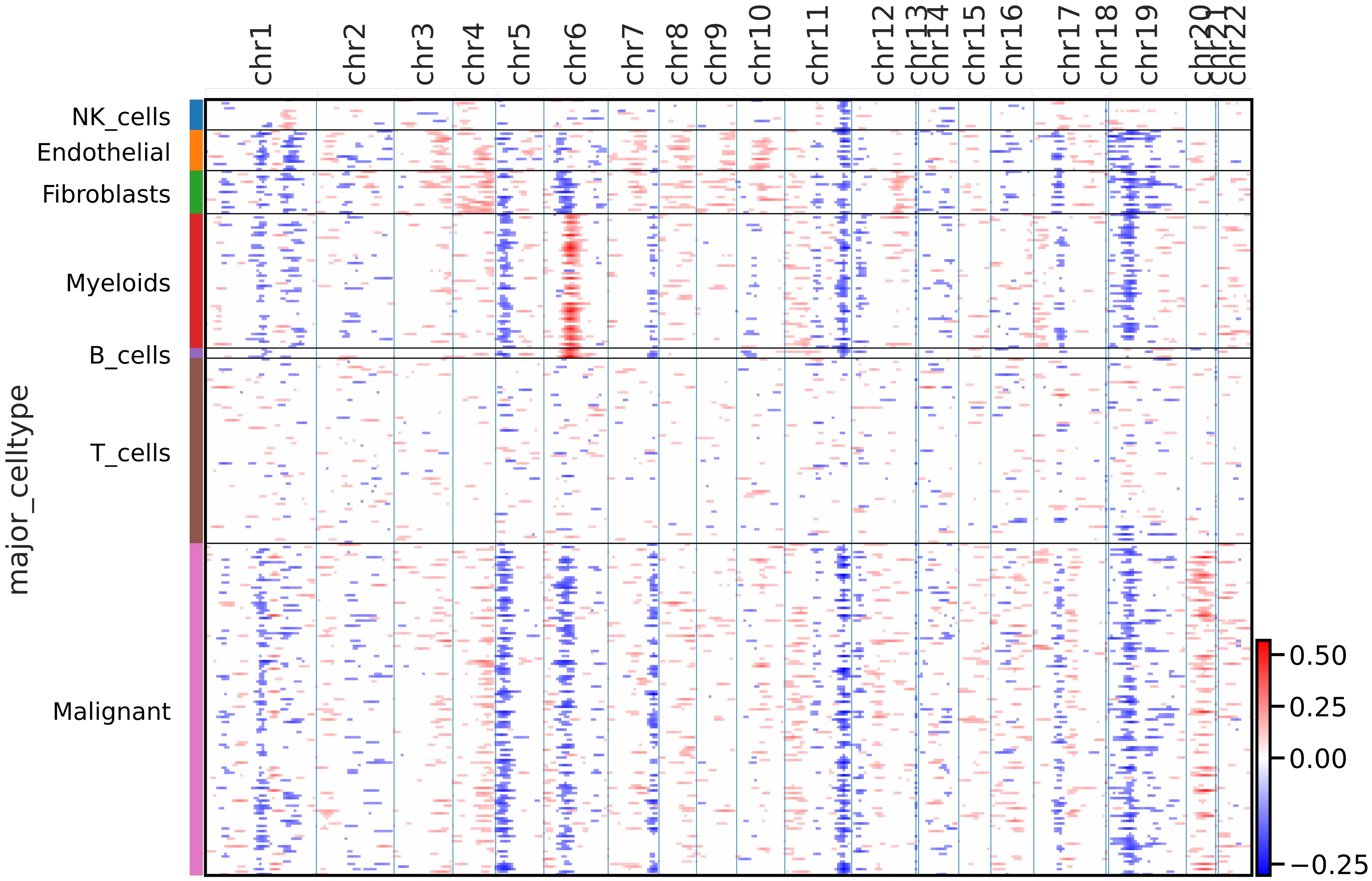

chr1
chr2
chr3
chr4
chr5
chr6
chr7
chr8
chr9
chr10
chr11
chr12
chr13
chr14
chr15
chr16
chr17
chr18
chr19
chr20
chr21
chr22
NK_cells
Endothelial
Fibroblasts
Myeloids
B_cells
T_cells
Malignant
major_celltype
0.50
0.25
0.00
−0.25

Primary 8

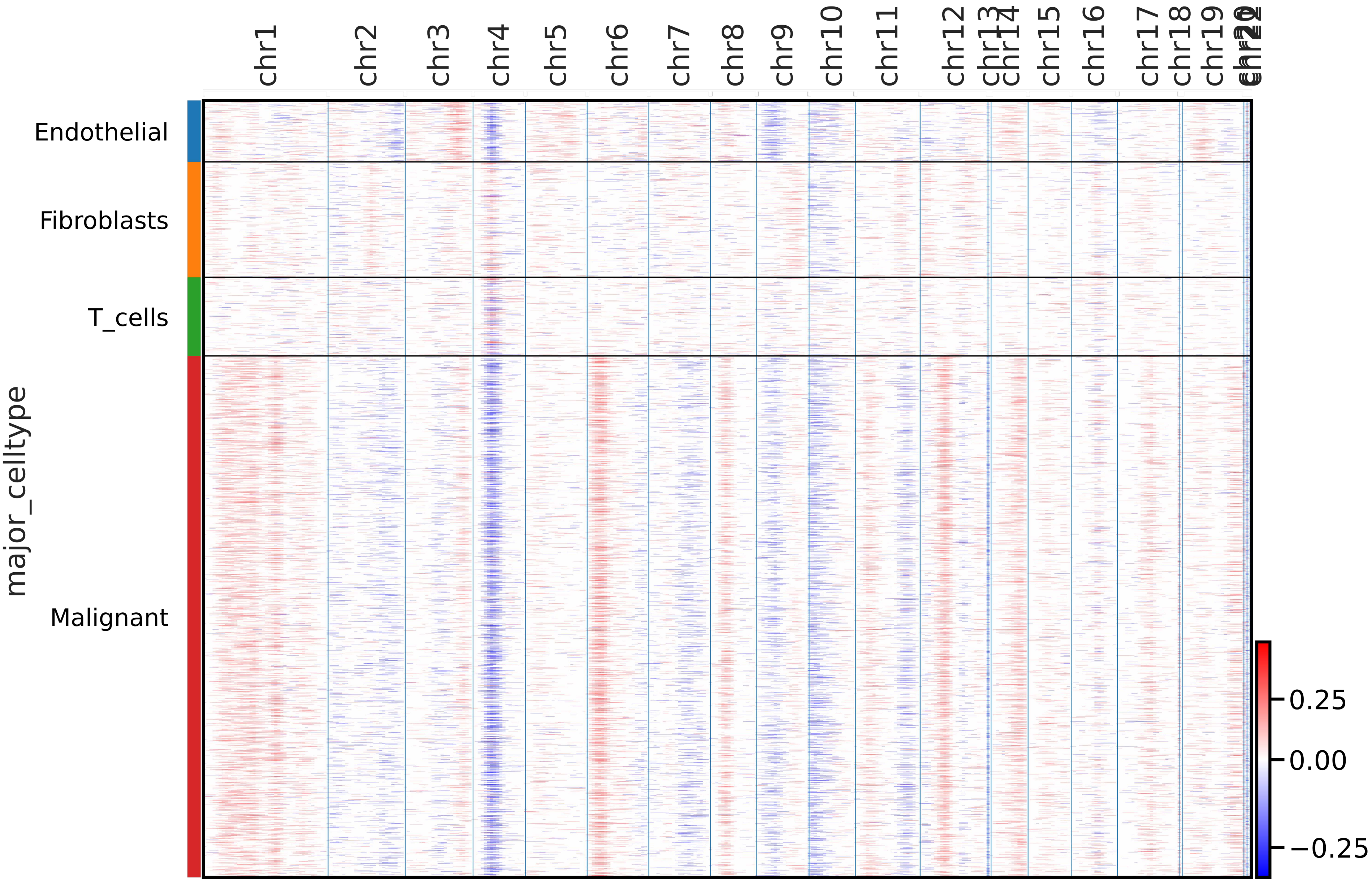
chr1
chr2
chr3
chr4
chr5
chr6
chr7
chr8
chr9
chr10
chr11
chr12
chr13
chr14
chr15
chr16
chr17
chr18
chr19
Endothelial
Fibroblasts
T_cells
Malignant
major_celltype
0.25
0.00
−0.25

Primary 9

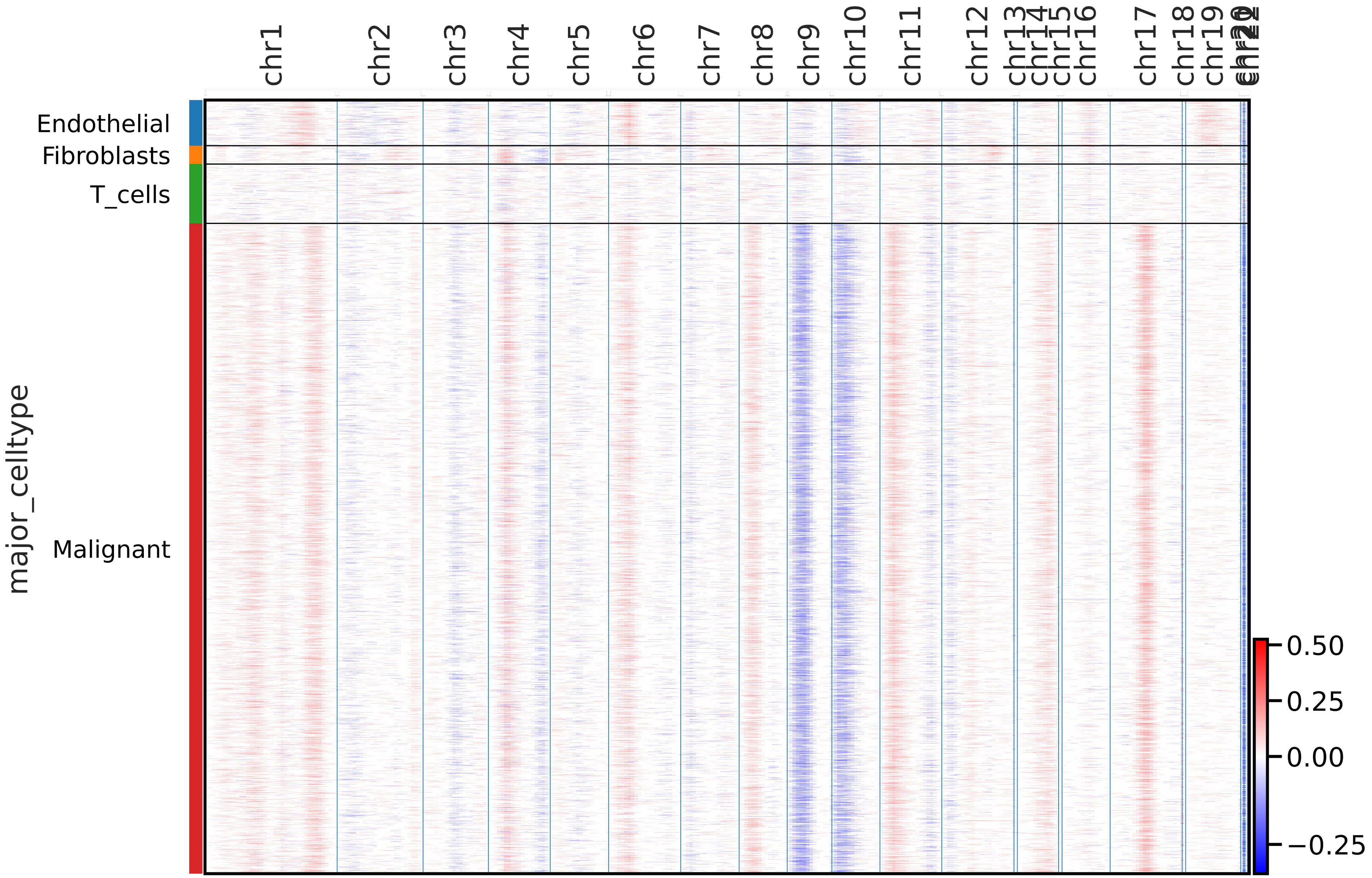

chr1
chr2
chr3
chr4
chr5
chr6
chr7
chr8
chr9
chr10
chr11
chr12
chr13
chr14
chr15
chr16
chr17
chr18
chr19
Endothelial
Fibroblasts
T_cells
Malignant
major_celltype
0.50
0.25
0.00
−0.25

Primary 10

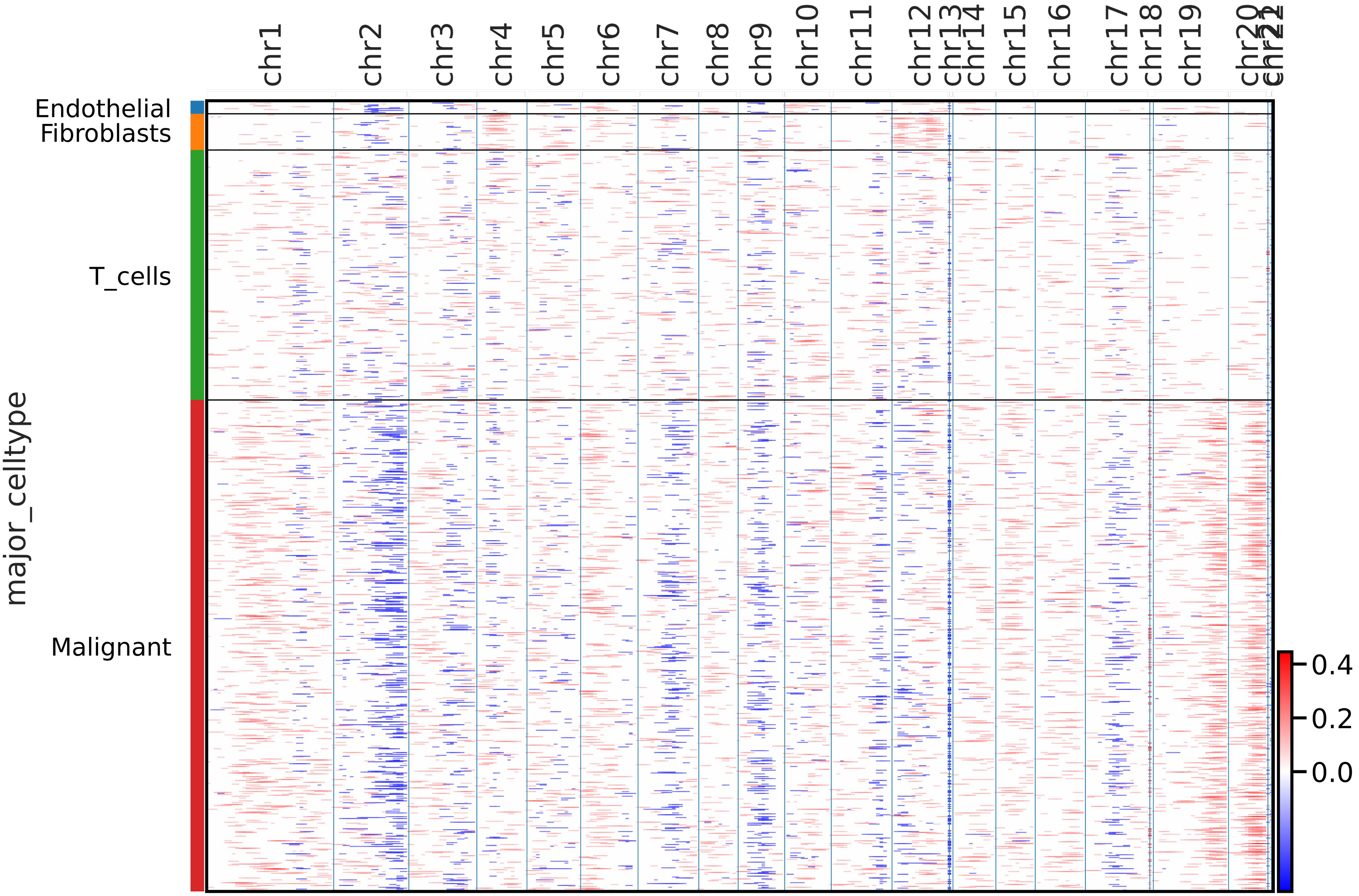

chr1
chr2
chr3
chr4
chr5
chr6
chr7
chr8
chr9
chr10
chr11
chr12
chr13
chr14
chr15
chr16
chr17
chr18
chr19
chr20
Endothelial
Fibroblasts
T_cells
Malignant
major_celltype
0.4
0.2
0.0

Primary 11

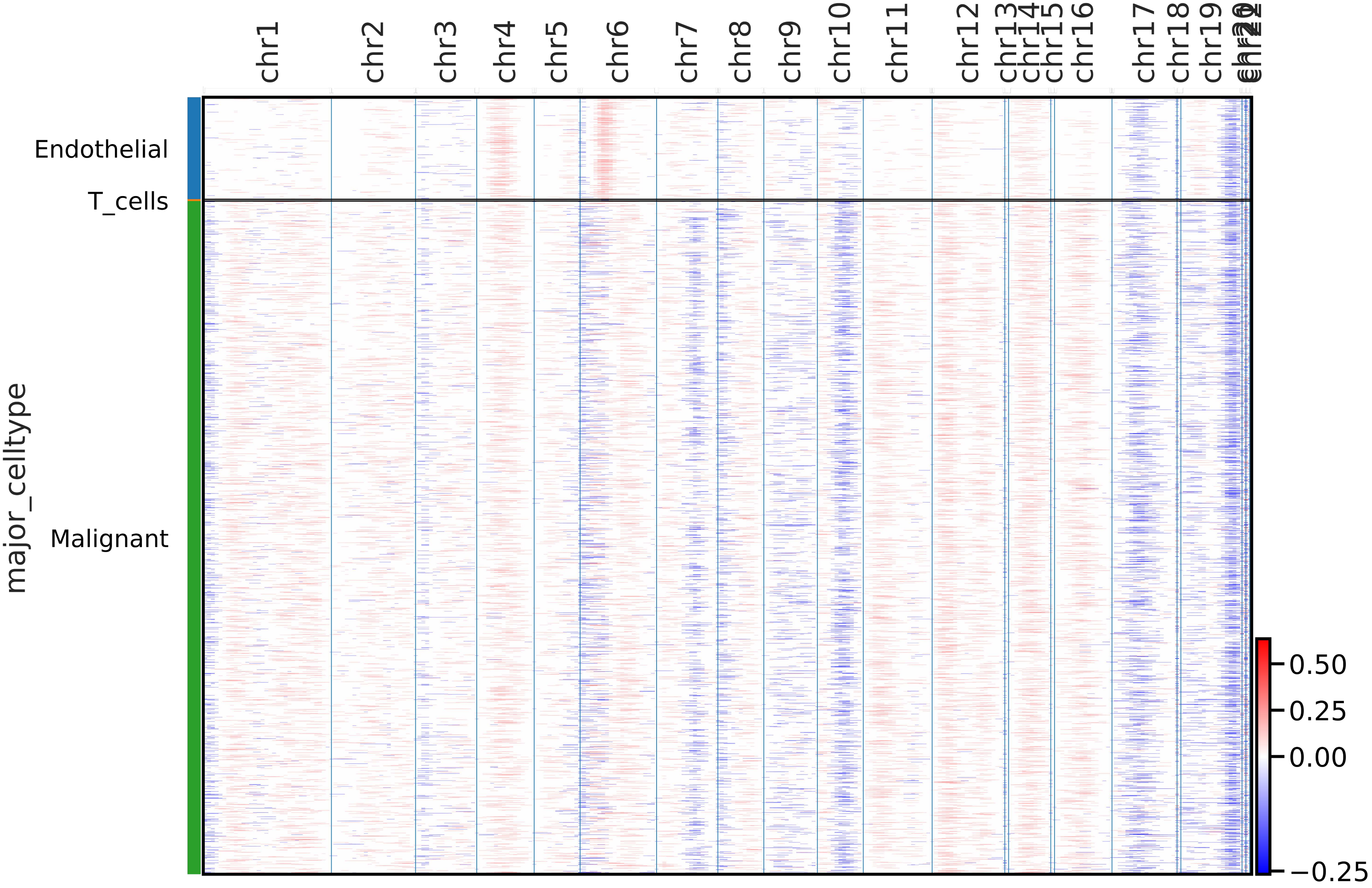
chr1
chr2
chr3
chr4
chr5
chr6
chr7
chr8
chr9
chr10
chr11
chr12
chr13
chr14
chr15
chr16
chr17
chr18
chr19
Endothelial
T_cells
Malignant
major_celltype
0.50
0.25
0.00
−0.25

Primary 12

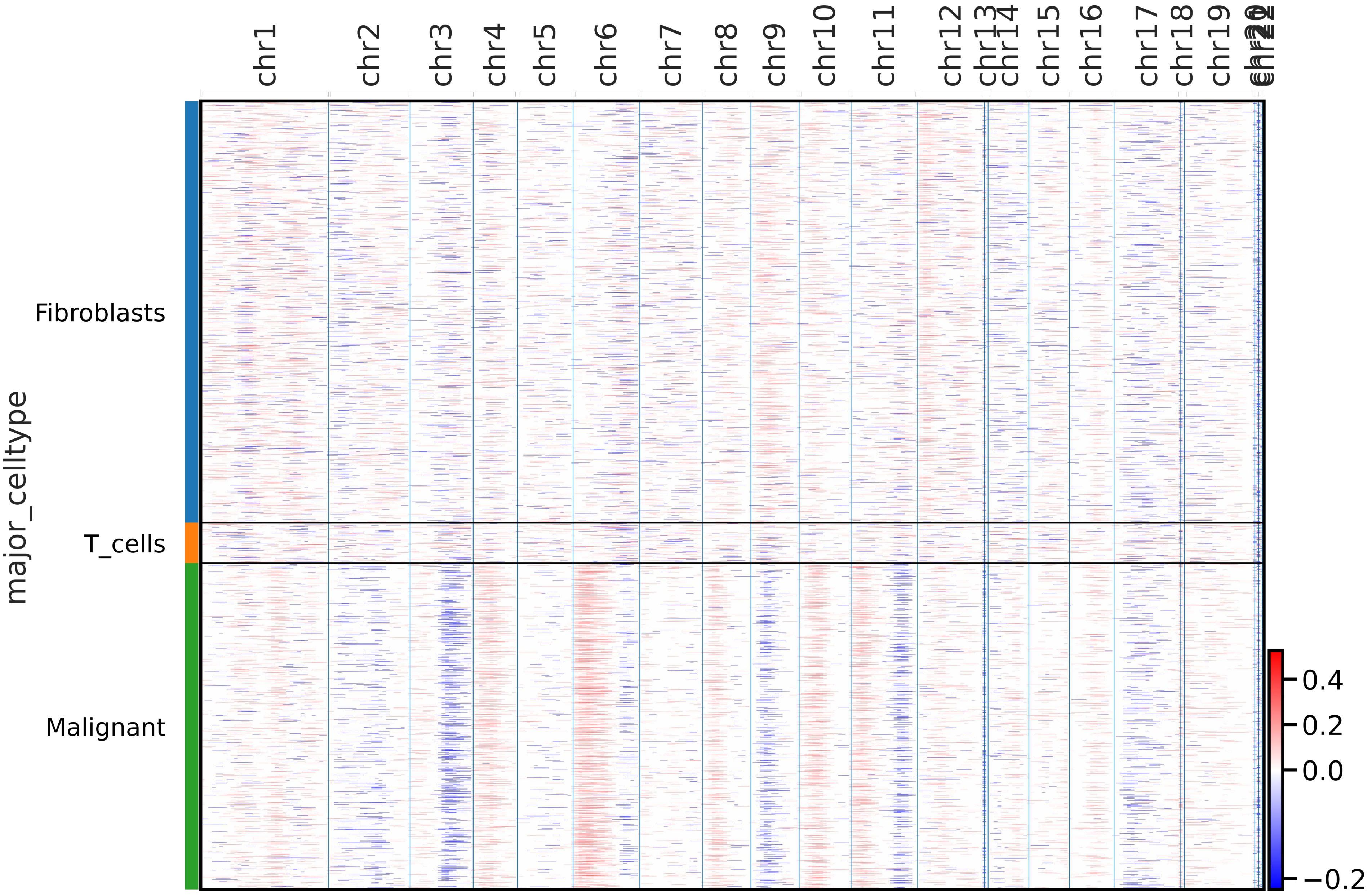
chr1
chr2
chr3
chr4
chr5
chr6
chr7
chr8
chr9
chr10
chr11
chr12
chr13
chr14
chr15
chr16
chr17
chr18
chr19
major_celltype
Fibroblasts
T_cells
Malignant
0.4
0.2
0.0
−0.2

Metastasis 1

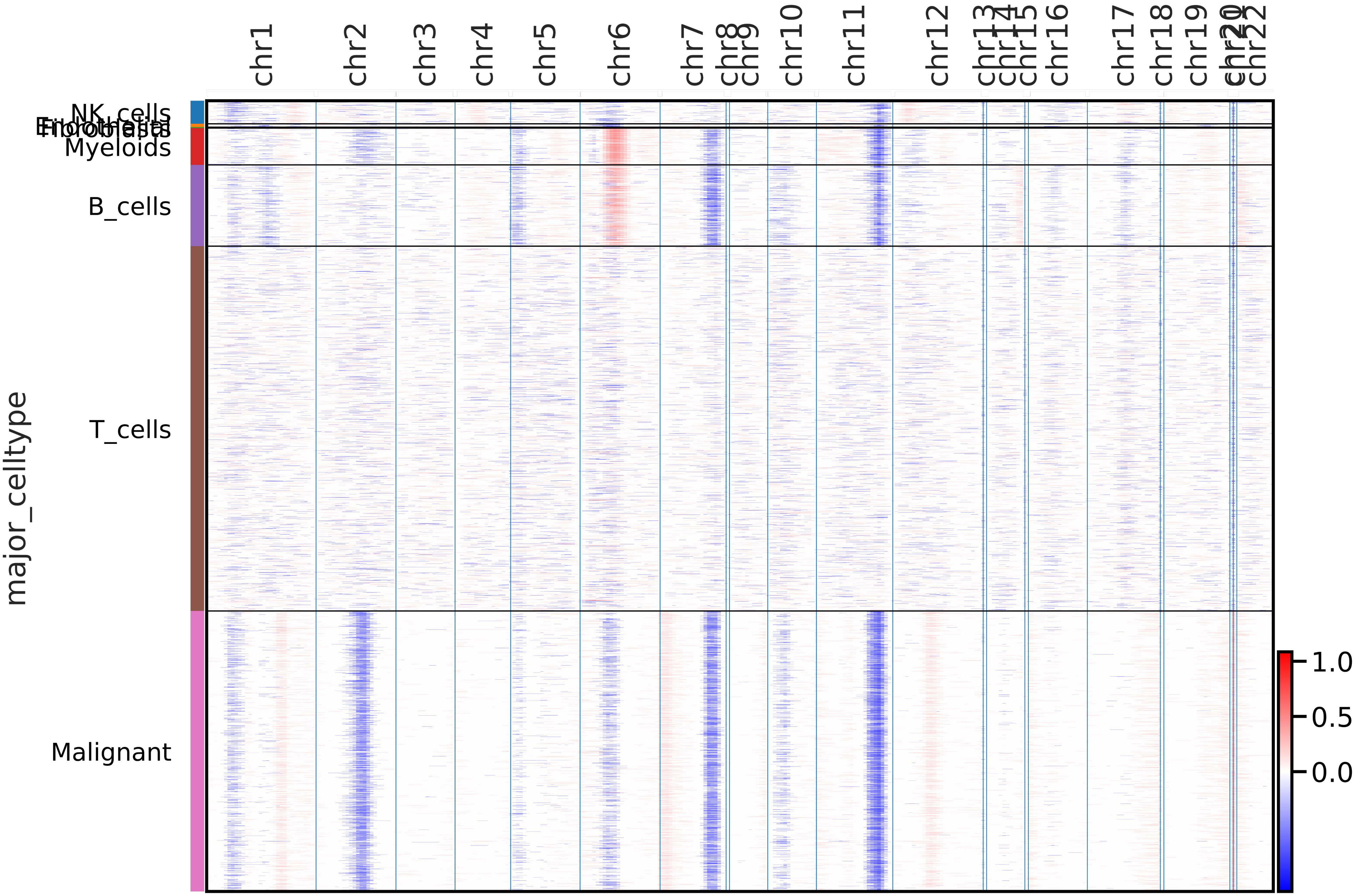

Metastasis 2

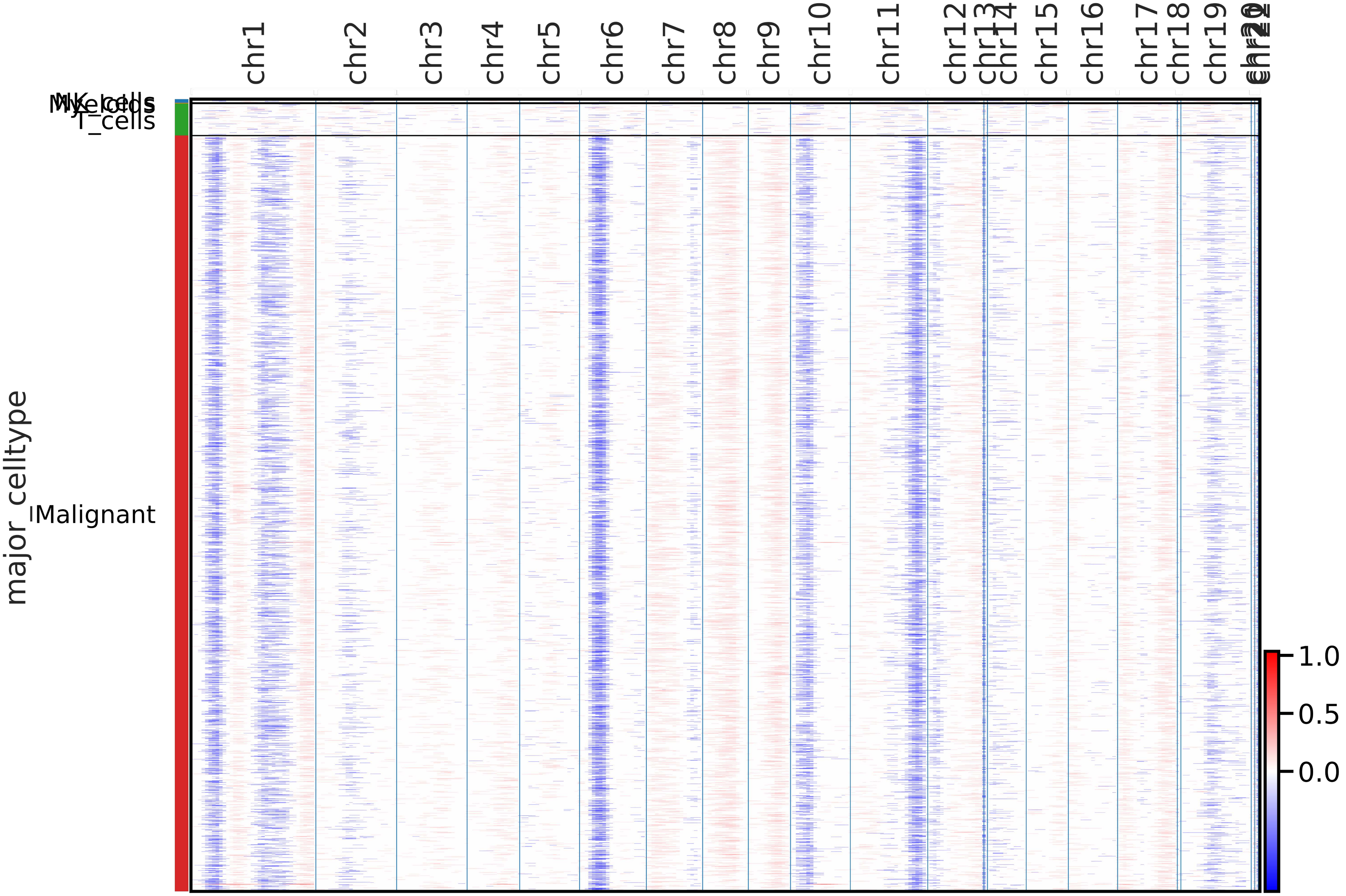

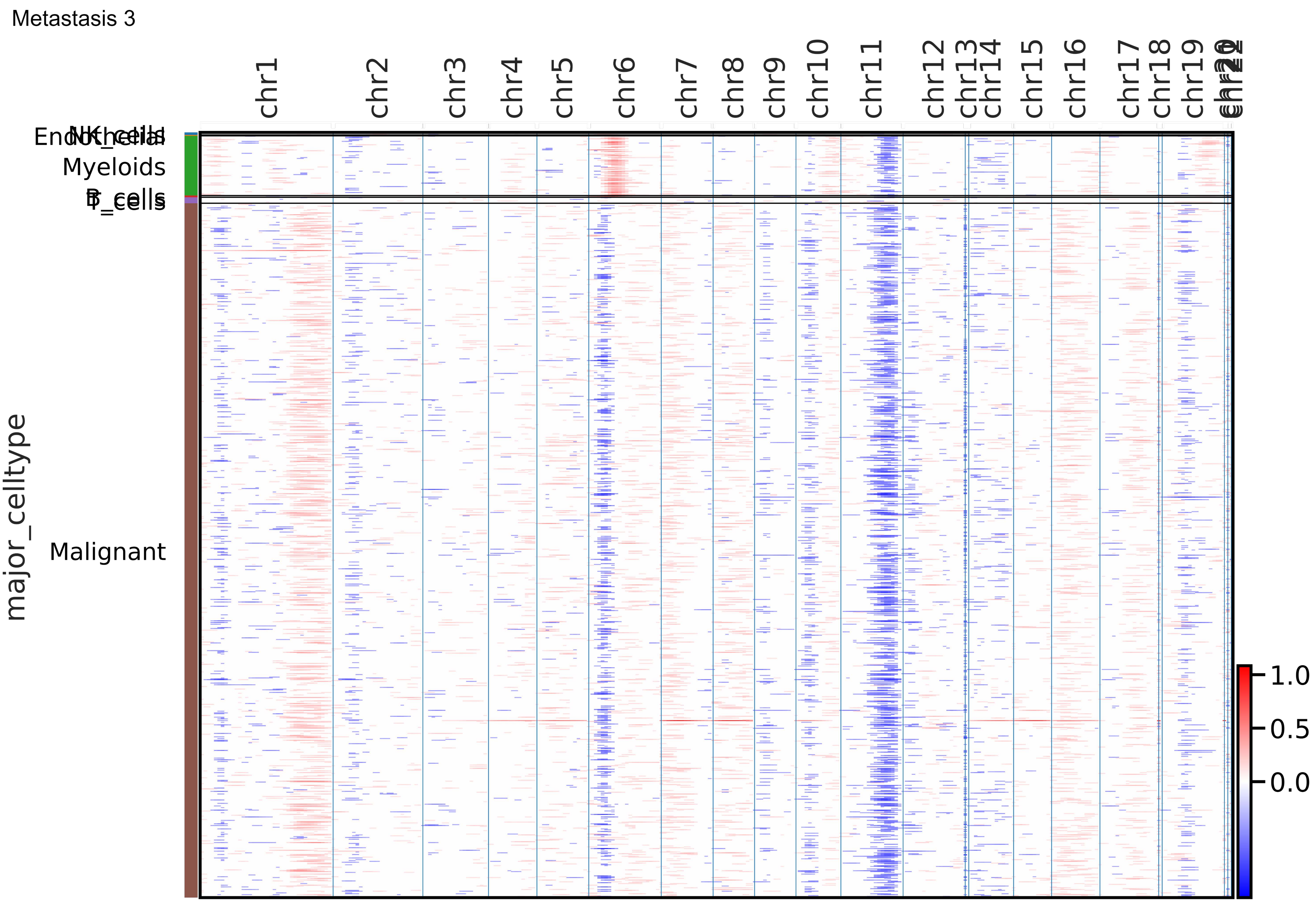

Metastasis 3
chr1
chr2
chr3
chr4
chr5
chr6
chr7
chr8
chr9
chr10
chr11
chr12
chr13
chr14
chr15
chr16
chr17
chr18
chr19
NK_cells
Endothelial
Myeloids
B_cells
T_cells
Malignant
major_celltype
1.0
0.5
0.0

Metastasis 4

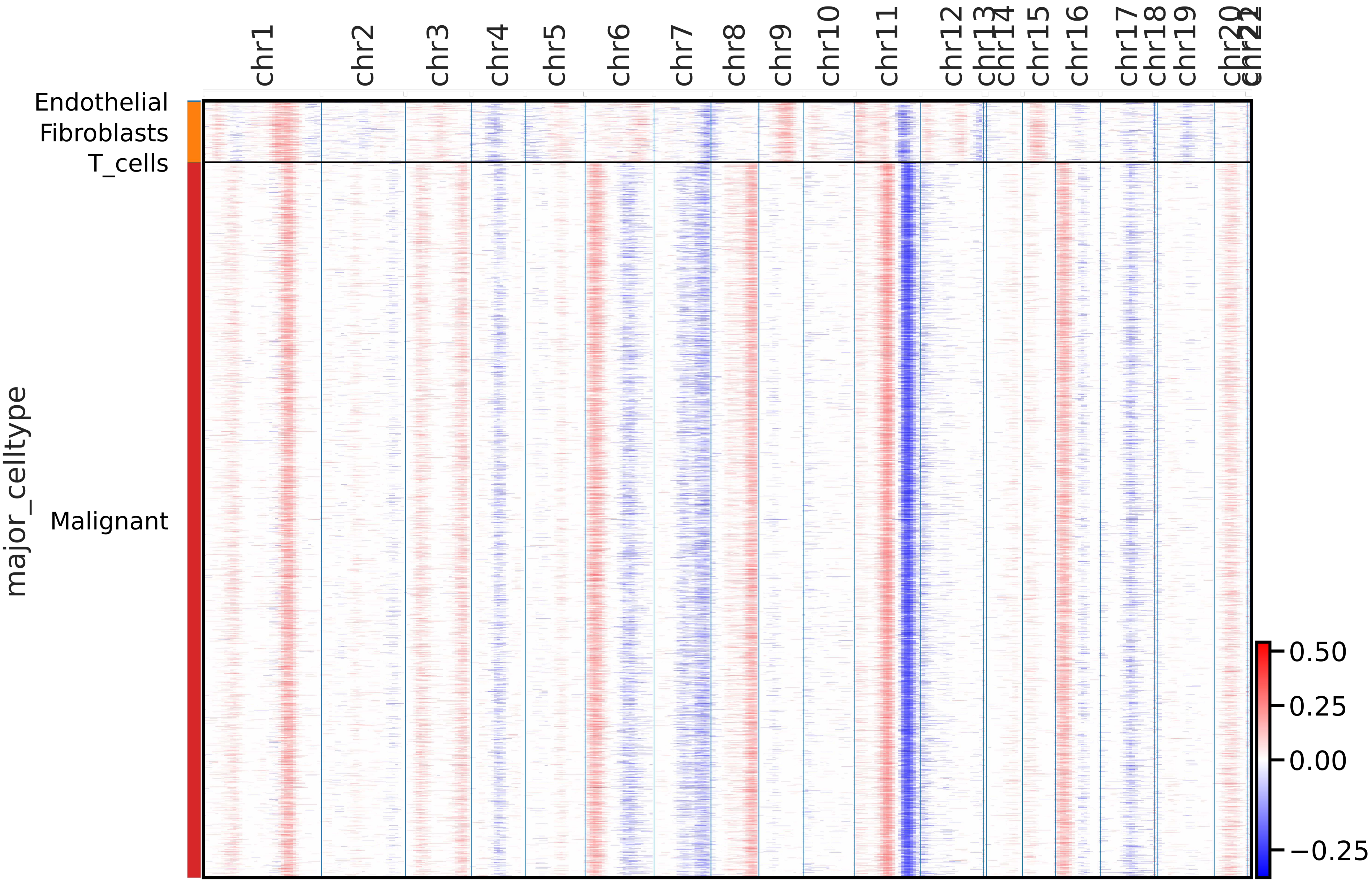

Metastasis 5

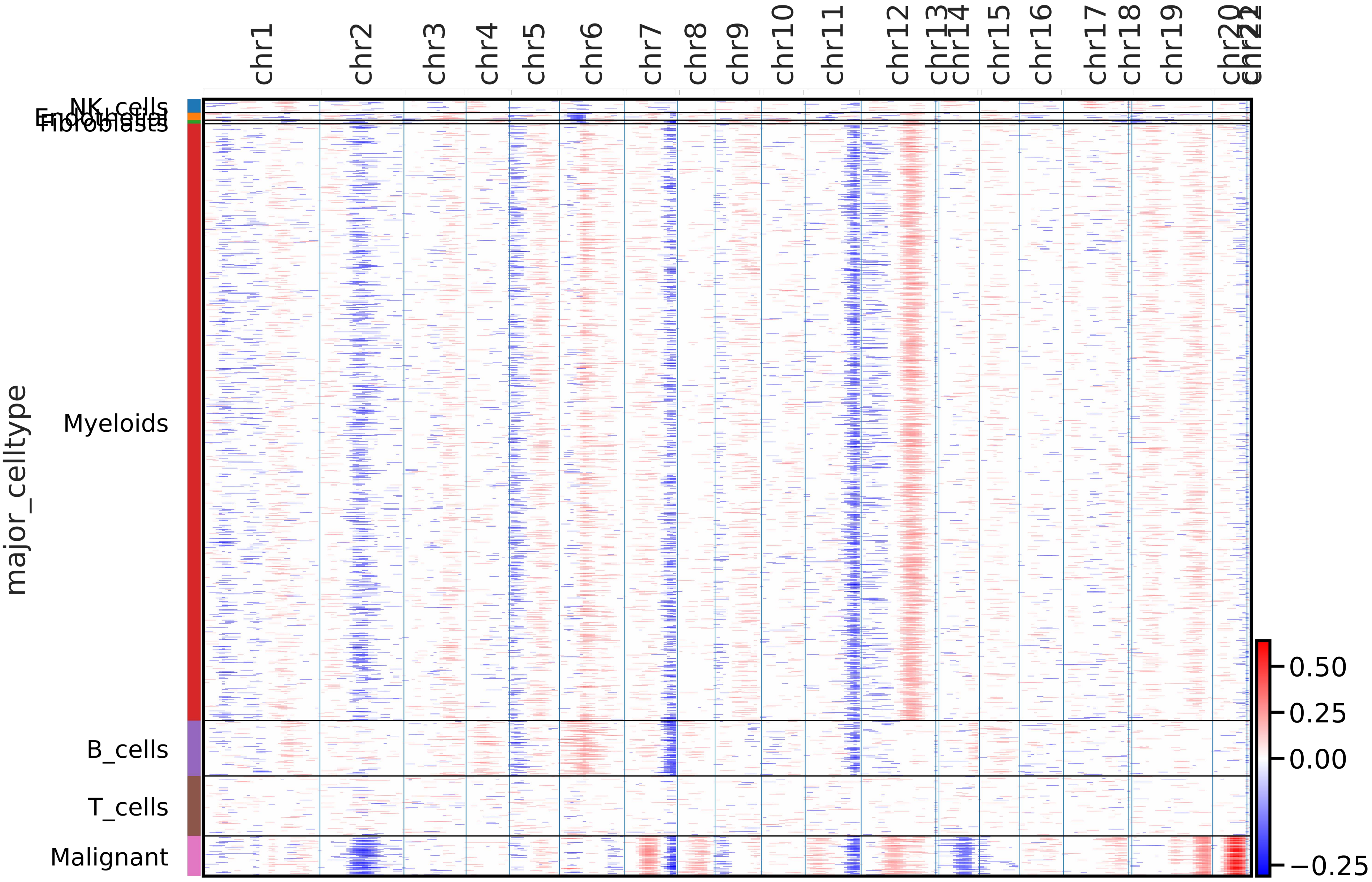

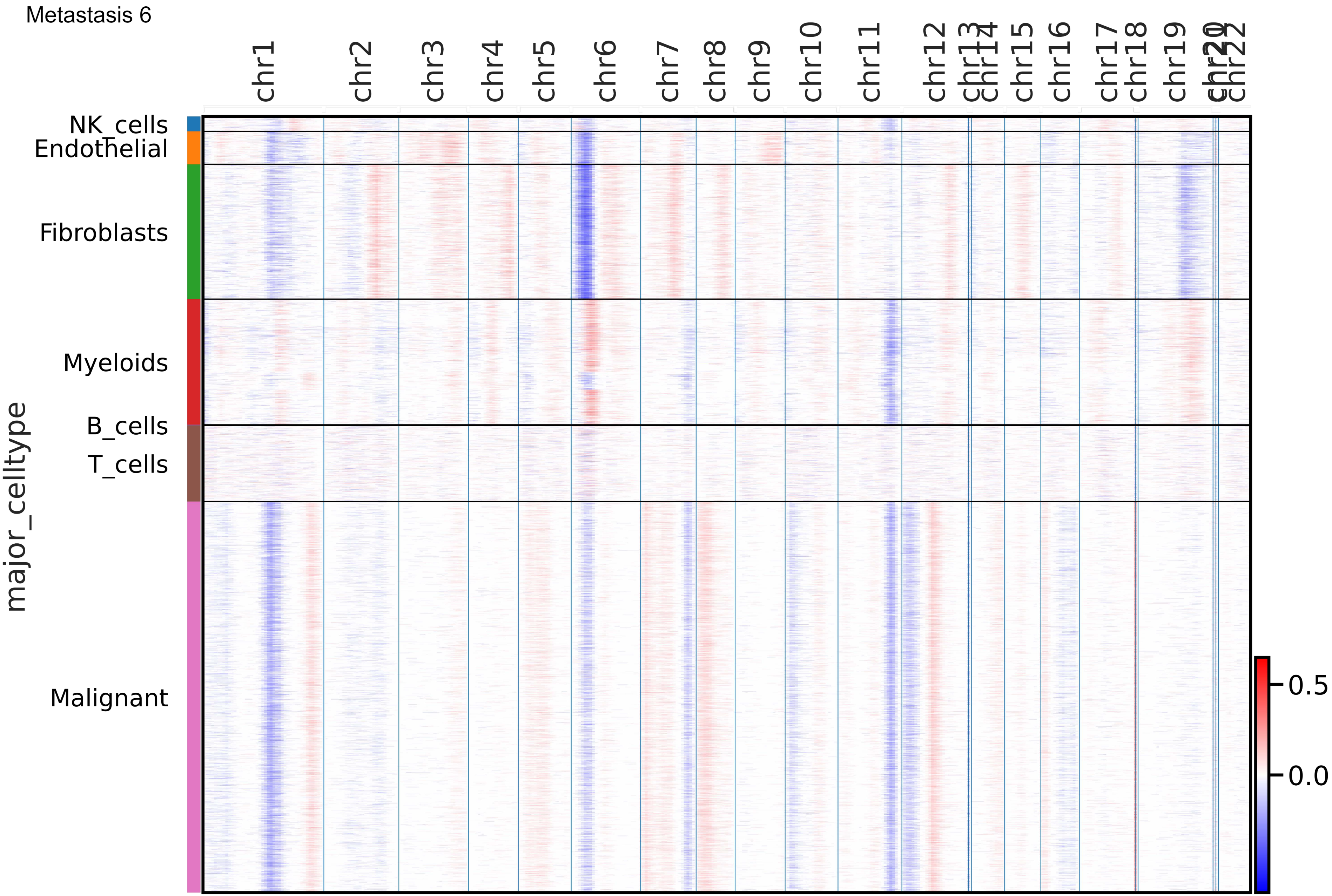
Metastasis 6
chr1
chr2
chr3
chr4
chr5
chr6
chr7
chr8
chr9
chr10
chr11
chr12
chr13
chr14
chr15
chr16
chr17
chr18
chr19
chr20
chr21
chr22
NK_cells
Endothelial
Fibroblasts
Myeloids
B_cells
T_cells
Malignant
major_celltype
0.5
0.0

Metastasis 7

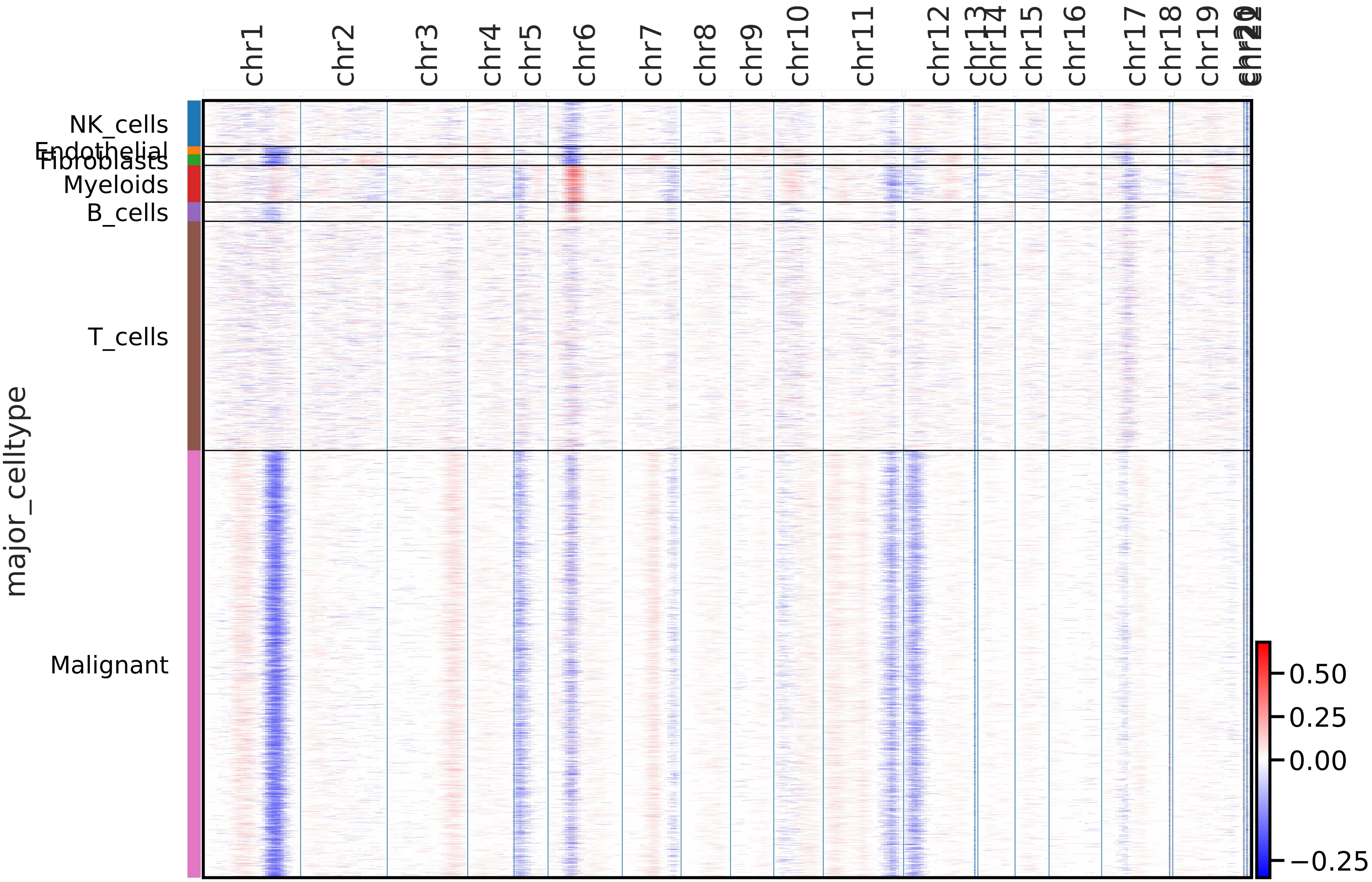

Metastasis 8

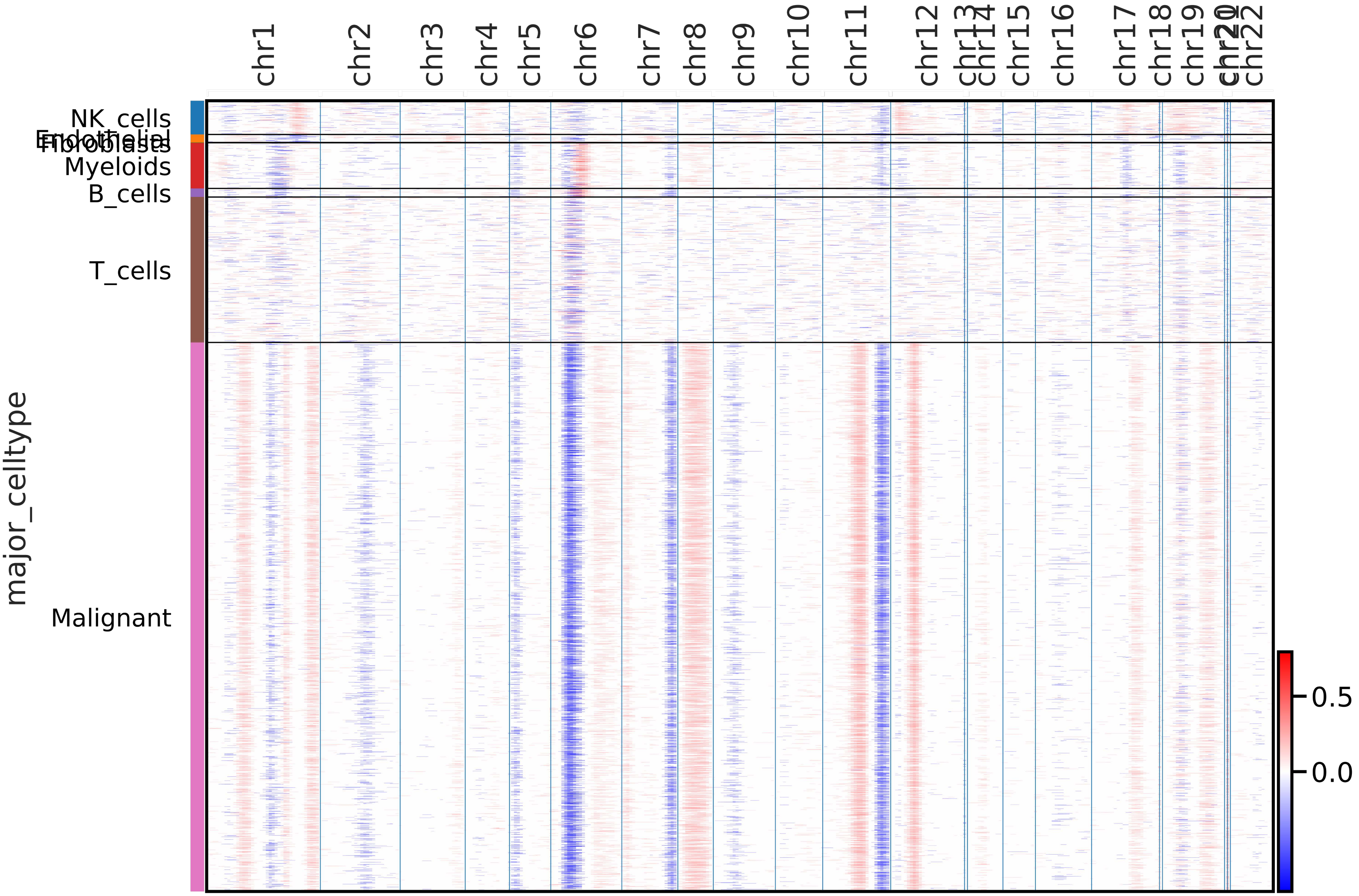

Metastasis 9

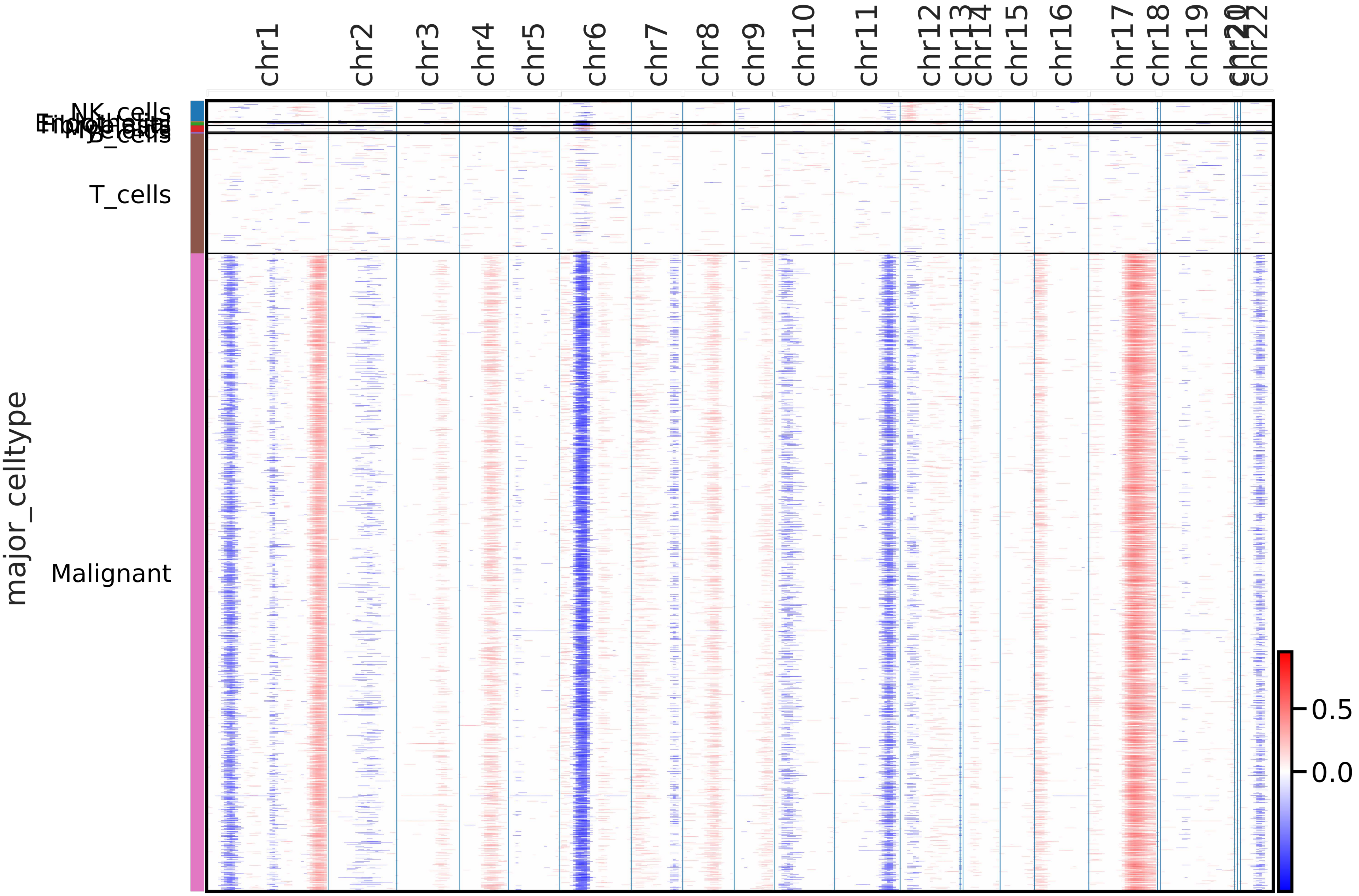

Metastasis 10

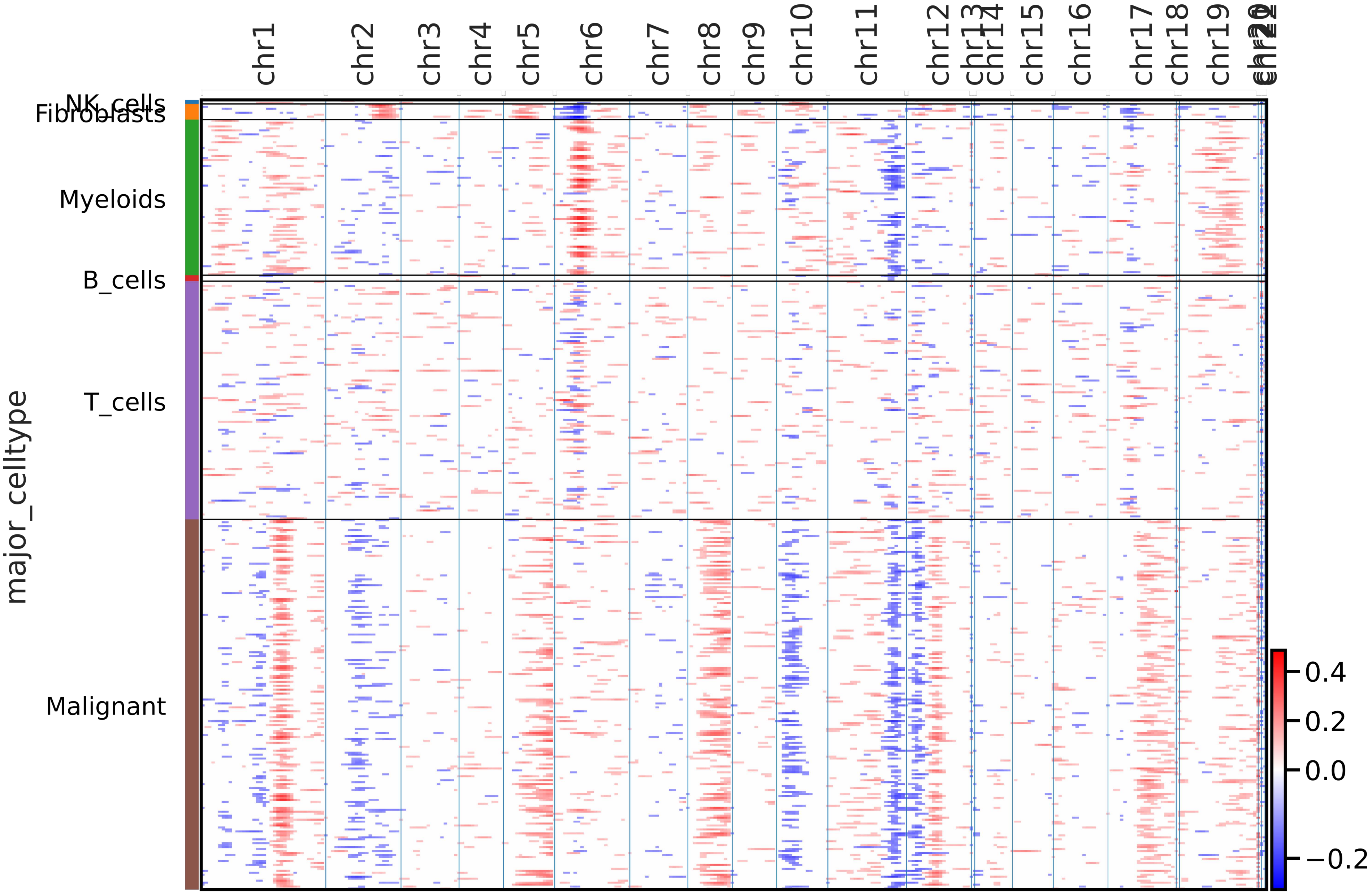

Metastasis 11

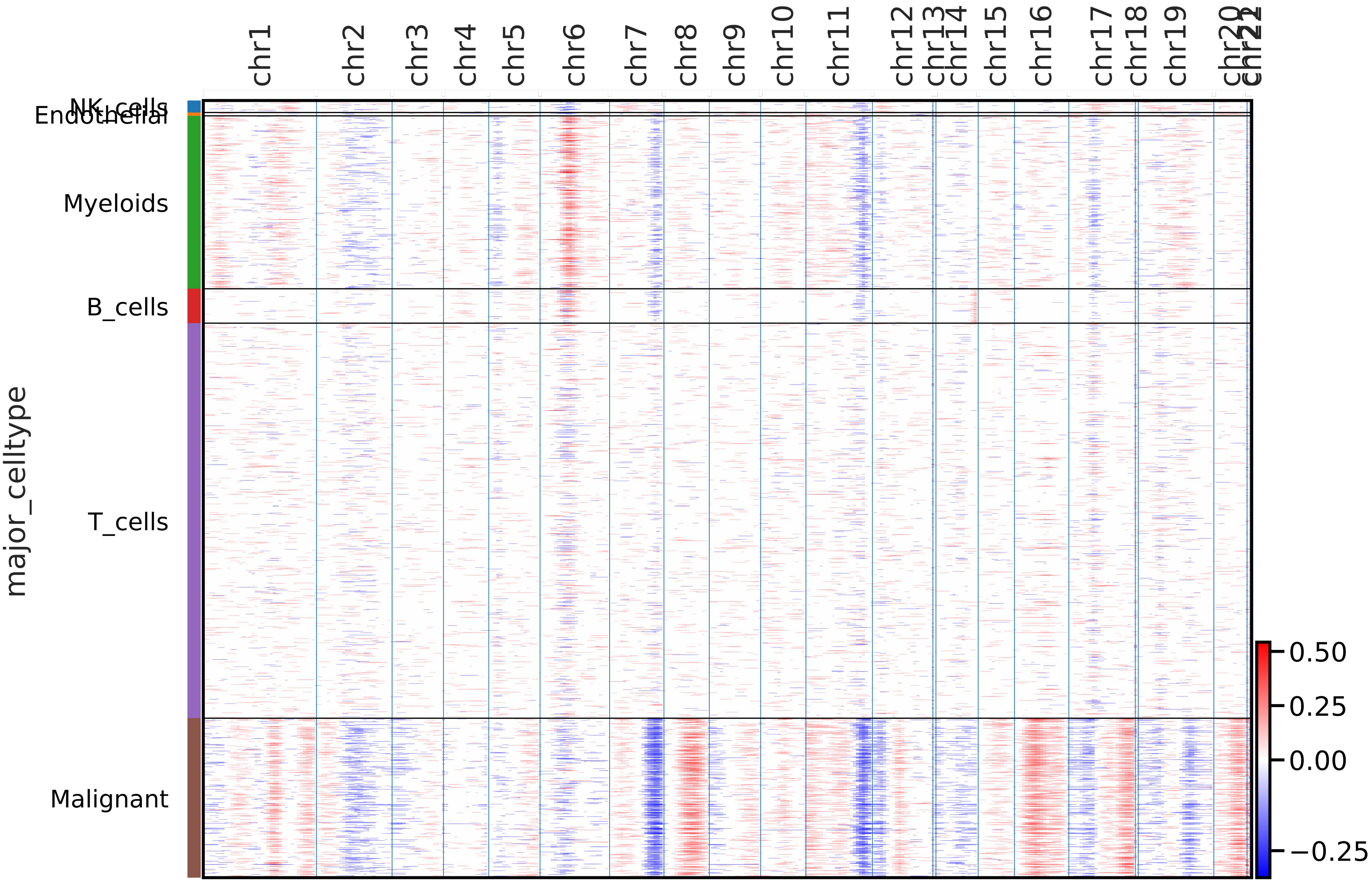
